# Gut microbiome activity predicts risk of type 2 diabetes and metformin control in a large human cohort

**DOI:** 10.1101/2021.08.13.21262051

**Authors:** Nan Shen, Nevenka Dimitrova, Cleo H. Ho, Pedro J. Torres, Francine R Camacho, Ying Cai, Momchilo Vuyisich, Damon Tanton, Guruduth Banavar

## Abstract

Recognizing and treating the early stages of type 2 diabetes (T2D) is the most cost effective way to decrease prevalence, before heart disease, renal disease, blindness, and limb amputation become inevitable. In this study, we employ high resolution gut microbiome metatranscriptomic analysis of stool samples from 53,970 individuals to identify predictive biomarkers of type 2 diabetes progression and potential for diagnosis and treatment response. The richness of the metatranscriptomic data enabled us to develop a T2D risk model to delineate individuals with glycemic dysregulation from those within normal glucose levels, with ROC-AUC of 0.83+/-0.04. This risk score can predict the probability of having insulin dysregulation before detecting high glycated hemoglobin (HbA1c), the standard-of-care marker for prediabetes and diabetes. Additionally, a machine learning model was able to distinguish novel metatranscriptomic features that segregate patients who receive metformin and are able to control their HbA1c from those who do not. These discoveries set the stage for developing multiple therapeutic avenues for prevention and treatment of T2D.

## Introduction

Type 2 diabetes (T2D) is the most prevalent endocrine disease, affecting more than 400 million people worldwide, and this number is expected to rise to 700 million by the year 2025 (Zhou et al., 2016) (Chatterjee et al., 2017). The increased prevalence of T2D is a major challenge for healthcare systems globally and novel tools are needed to reduce the burden of T2D. Recognizing and treating the early stages (and even predisease) is the most cost effective way to treat T2D, before end-organ damage, such as heart disease, stroke, renal disease, blindness and limb amputation, ensues. (Bailes, 2002). Multiple genomic and transcriptomic studies have been conducted on the analysis of human host samples and animal models to elucidate the molecular mechanisms of disease progression (Lawlor et al., 2017; Segerstolpe et al., 2016; Sengupta et al., 2009). Although it is a complex disorder influenced by both genetic and environmental components, central obesity (visceral fat) is known to be the driving risk factor (Hu, 2011).

In recent years, the gut microbiome has emerged as a prominent new frontier in medical research, having been found to be implicated in a range of chronic and acute health conditions and outcomes. Gut microbiome research has primarily focused on genomic DNA. Multiple studies have shown that the gut microbiome is altered in individuals with metabolic disorders, such as obesity (Turnbaugh et al., 2009) (Ley et al., 2006), T2D (Larsen et al., 2010) (Gurung et al., 2020) and progression of glucose intolerance (Zhang et al., 2013), and that the gut microbiome may be a causal factor in the development of these disease processes. (Wang et al., 2020). (Ridaura et al., 2013). Additionally, research has evaluated the predictive power of the gut microbiome for early T2D risk detection (Li et al., 2020). Findings in the existing metagenomic studies, indicated that *Ruminococcus, Fusobacterium, Blautia* are positively associated, while *Bifidobacterium, Bacteroides, Faecalibacterium, Akkermansia* and *Roseburia* are negatively associated with type 2 diabetes (Gurung et al., 2020).

While previous metagenomic studies have revealed compelling insights into the role that microorganisms play in the development of T2D, these approaches have described the presence of organisms and genes, rather than activity of these microbiome elements. Using non-invasive fecal samples, Viome’s clinical grade (CLIA License Number: 32D2156145) and fully automated metatranscriptomic technology (Hatch et al., 2019) has the potential to facilitate large-scale public health applications, including early diagnosis and risk assessment for T2D.

In this study, we utilized the gut microbiome of 53,970 individuals, using a gut metatranscriptome approach combined with multivariate and machine learning analyses, in an effort to answer several questions:

1. How does the microbiome change as an individual progresses from non-diabetes (normal glucose homeostasis), to prediabetes, to T2D? And, more importantly, what are the microbiome functional changes that take place throughout this spectrum of disease? In this paper, we delineate the microbiome differences between individuals with non-diabetes and prediabetes and we compare these changes with the differences between the microbiomes of individuals with prediabetes and T2D. By establishing taxonomic and functional progression, we should be able to inform further target discovery for modulating the pathways of microbiome with medications, prebiotic, probiotic, or other supplement recommendations.
2. Can the gut microbiome provide insight into the early diagnosis of either prediabetes or T2D? With advanced predictive modeling, we aimed to develop a metabolic disease risk score, based on the gut metatranscriptome, that can predict the probability of having insulin dysregulation before detecting high glycated hemoglobin (HbA1c), which is used as the standard-of-care diagnostic for prediabetes and diabetes conditions. By using this risk score for T2D, patients can then take action to either prevent or postpone further progression of disease. Our aim is to pick up early signals for the beginning of metabolic disease, not just the beginning of diagnosis of the same. Related question is to understand if people who follow food recommendations would show improved microbiome diabetes risk scores?
3. What are the microbiome correlates of response to current treatments for individuals with T2D? There are multiple medications that are used in the treatment of T2D. For individuals who have already been diagnosed with T2D (or those with a prediabetes condition receiving treatment in the form of medication), can we identify microbiome-related determinants of response to treatment? Most frequently, patients receive metformin, and we aim to determine the microbiome signature of controlling HbA1c while they are taking metformin. For the patients who are not able to control their HBA1c while on metformin, this signature may point to additional interventions that can assist with glucose control.
4. For individuals who choose not to take treatment in the form of medications or insulin therapy (and instead rely on diet and lifestyle changes), can we determine if there are microbiome correlates of response to these non-medication treatments? Can these correlates inform how to modulate the microbiome with a positive treatment effect. In other words, are there microbiome pathways that can be modulated so as to achieve better glycemic control? If so, then regular surveillance of these changes should be aided by the microbiome readout over time.

Table 1 shows the characteristics of our discovery (n=50,942) and validation (n=3,028) cohorts.

**Table 1:** Study cohorts.

| **Table 1: Study cohorts** |  |  |
| --- | --- | --- |
|  | Discovery | Validation |
| Number of samples | 50,942 | 3,028 |
| Non-diabetes controls | 50,251 | 2,000 |
| Pre-diabetes | 462 | 770 |
| Type II Diabetes | 229 | 258 |
| Sex (% female) | 58.7 | 65.75 |
| Non-diabetes controls | 58.6 | 63.25 |
| Pre-diabetes | 68.2 | 75.19 |
| Type II Diabetes | 64.4 | 56.98 |
| Age (y) mean ± std | 41.9 ± 13.7 | 46.6 ± 15.1 |
| Non-diabetes controls | 41.8 ± 13.7 | 41.3 ± 13.6 |
| Pre-diabetes | 57.3 ± 11.4 | 55.6 ± 12.5 |
| Type II Diabetes | 59.9 ± 11.4 | 60.7 ± 11.2 |
| BMI (all / non-med) |  |  |
| Normal < 25 | 29949 / 17159 | 1442 |
| Overweight 25 to <30 | 14439 / 8013 | 880 |
| Obese ≥ 30 | 5863 / 2810 | 706 |

## Results

### Phenotype stratification

Figure 1 describes the clinical phenotypes based on the self-reported labels in our study. The non-diabetic group includes presumed healthy individuals who are not reporting medical or mental conditions and are not actively taking medications, as well as individuals who have reported comorbidities or are taking medications. The non-diabetic group excludes anyone taking antibiotics within one month of sample collection, proton pump inhibitors and/or acid suppressants, had abdominal surgery or report diseases that may affect gut microbiome significantly (e.g. IBS, IBD, colon cancer). For this group we have matched the age, sex and body mass index (BMI) to those in the prediabetic and T2D groups.

**Figure 1.**
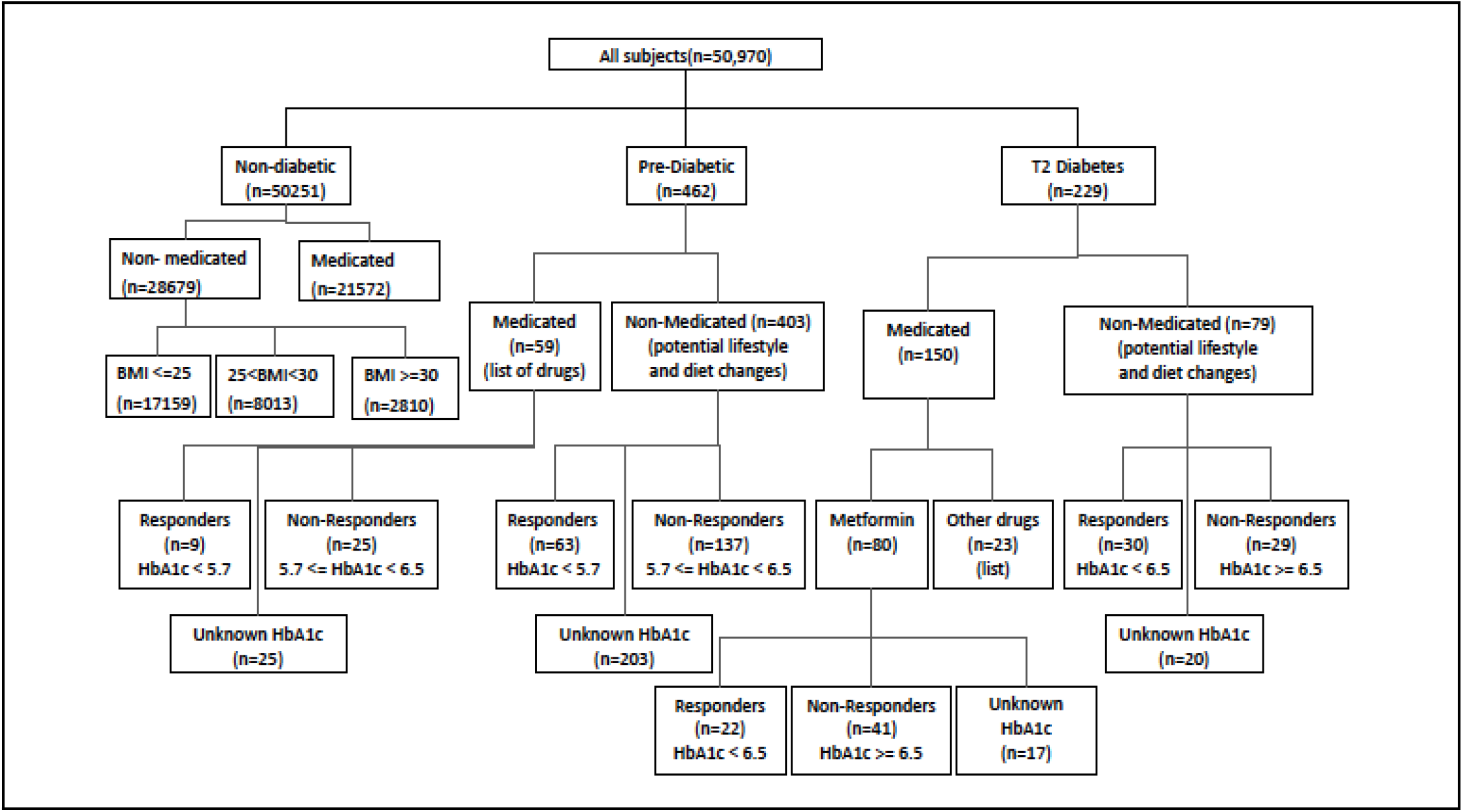
Discovery cohort, used for statistical analysis and model training/development. (Note: Metformin (n=80) are people using only metformin; other drugs (n=23) are people using drugs other than metformin; the remainder of medicated (n=150) were people using metformin along with other drugs, and are not shown here, nor were used in any analysis.)

The first stratification is based on the type of diagnosis that patients report, i.e., prediabetes and T2D. Within these two groups, there are individuals who received treatment in the form of medications, as well as individuals who are not on medications, and instead have diet and lifestyle changes. We consider within the T2D group that subjects who have reported HbA1c <6.5 to be able to control their HbA1c using medication, and the subjects with >=6.5 to not be able to control their HbA1c. We used the stratification in Figure 1 to analyze disease progression, diagnosis and medication control, using multivariate statistical analysis and machine learning.

### Summary of analyses

Table 2 presents a summary of the comparison cohorts and relevant analyses related to disease progression and T2D risk score. The first row represents the main cohort comparison used as the basis for the T2D risk score, to be described in more detail below. The next three rows represent the analyses for disease progression. The last row presents the analysis for the obesity/BMI related microbial features.

**Table 2.** Summary of methods and results for progression of disease and diagnostics for type 2 diabetes (T2D) *med, medicated; non-med, non medicated

| Comparison cohorts | Cohort matching criteria | Diff. Prev. sig. feats. (p.adj $< 0.05$) | Diff. Expr. sig. feats. (p.adj $< 0.05$) | Classific ation method | Mean ROC-AUC | Mean balanced accuracy | Classifier #features (KOs species) + |
| --- | --- | --- | --- | --- | --- | --- | --- |
| T2D med+non-med* (n=206) vs Non-T2D med+non-med (n=50251) | age, sex, bmi | 1415 KOs<br>266 species<br>(Chi2 test) | 1352 KOs<br>338 species<br>(KW test) | SVM + effect size feature selection | 0.83 | 0.76 | 922 |
| T2D non-med (n=73) vs Non-T2D non-med (n=28679) | age, sex, bmi | 115 KOs<br>78 species<br>(Chi2 test) | 206 KOs<br>111 species<br>(KW test) | SVM + effect size feature selection | 0.82 | 0.73 | 922 |
| T2D non-med (n=73) vs Pre-T2D non-med (n = 129) | none | 0 KOs<br>0 species<br>(Chi2 test) | 1 KO<br>0 species<br>(KW test) | SVM + effect size feature selection | 0.61 | 0.56 | 695 |
| Pre-T2D non-med (n=129) vs Non-T2D non-med (n=28679) | age, sex, bmi | 677 KOs<br>97 species<br>(Chi2 test) | 546 KOs<br>75 species<br>(KW test) | SVM + effect size feature selection | 0.72 | 0.67 | 922 |
| BMI model for non-med: Normal $< 25$ (n=17159), Overweight 25 to $< 30$ (n=8013), Obese $\geq 30$ (n=2810) | none | NA | 5485 KOs<br>680 species<br>(KW test) | BMI features are incorporated into all other analyses | | | |

### Obesity-related microbial composition and functional relevance

One of the most frequently reported confounding factors for T2D disease progression is obesity. Here we first assessed the contribution of obesity phenotype to microbiome by applying a statistical Kruskall-Wallis (KW) model from 27,982 subjects in our cohort who had low BMI <25 (n=17159) and belong to a normal range, intermediate BMI>=25 and BMI <30 (n=8013) who belong to the overweight range and high BMI >=30 (n=2810) who belong to the obese range.

We observed that 5485 KOs and 680 species were statistically significant in a three-way KW analysis with FDR <0.05. In all our further analyses, we highlight the observed obesity and non-obesity-related features so that we can delineate the unique microbiome associations of the type 2 diabetes state.

### Disease Progression Descriptive statistics

In total, 4380 taxa and 6797 functional KOs were identified in our cohort. Figure 2 presents the differential expression, for the most statistically significant features (using Kruskal-Wallis method after Benjamini-Hochberg FDR correction, p-value<=0.05). Each figure presents non-T2D patients (in blue) who have not received any medication vs. T2D or prediabetes patients (in red). Supplementary figure s1 presents the differential prevalence corresponding to the same comparison groups.

**Figure 2:**
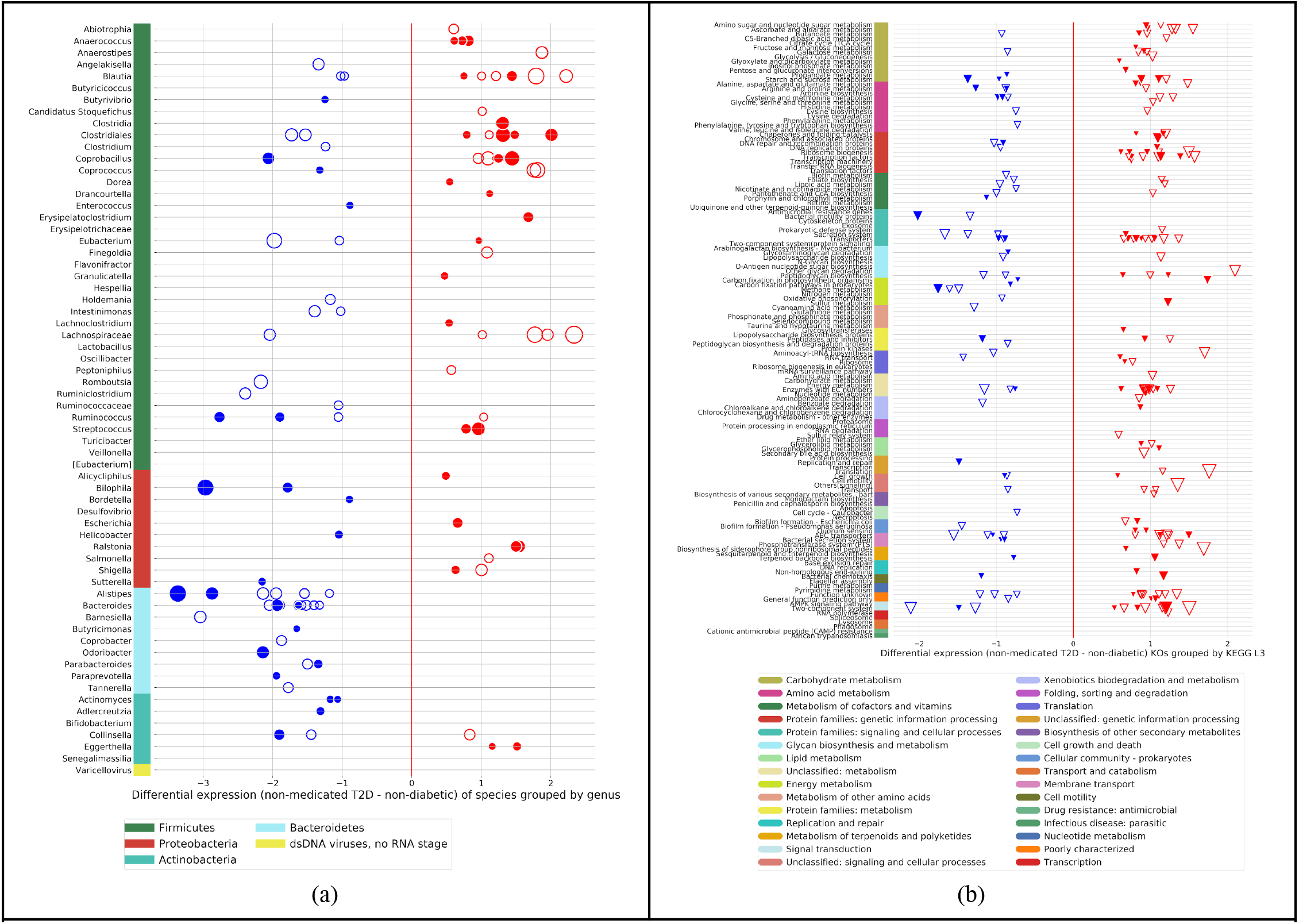
Progression of disease: differentially expressed features in a color-bar scatter plot (KW statistics, FDR p-value < 0.05). Panels present scatter plots of resulting patient features which include (a) differentially expressed species shown as circles between non-medicated T2D vs non-medicated non-T2D and (b) differentially expressed KOs shown as triangles between non-medicated T2D vs non-medicated non-T2D. Filled markers represent obesity related features, and hollow markers represent T2D-unique features.

Shown in scatter plots : we observe 75 out of 4380 unique differentially expressed microorganisms when we compare pre-diabetic vs. non-diabetic (see Supplementary Figure s1b), and we observe 111 unique microorganisms when we compare T2D vs non-diabetic samples (see Figure 2a). We observe 546 differentially expressed KOs in pre-diabetic vs. non-diabetic (see Supplementary figure s1d) and 206 differentially expressed KOs when we compare T2D vs non-diabetic subjects (see Figure 2b). In contrast with previous studies which are performed using 16s technology, we are able to observe a wider spectrum of differential expression and infer functional modules directly from the experimental observations.

Shown in side-by-side scatter plots (see Supplementary Figure s1): we observe 97 statistically significant species out of 4380 unique active microorganisms when we compare pre-diabetic vs. non-diabetic (see Supplementary Figure s1a - left panel), and we observe 78 unique microbial species when we compare T2D samples vs. non-diabetic (see Supplementary Figure s1a - right panel) using Chi-squared tests after Benjamini-Hochberg FDR correction, p-value<=0.05). In addition, the results from the SVM model are presented in Supplementary Figure s5.

We observe 677 prevalent KOs pre-diabetic vs. non-diabetic (see Figure s1c left panel) and 115 prevalent KOs when we compare T2D vs non-diabetic samples (see Figure s1c right panel).

### Ruminococcus and Blautia genera

We single out results from differential prevalence and expression of bacteria belonging to *Blautia* and *Ruminococcus*, genera which are reported in the literature as positively associated with type 2 diabetes. However, in our metatranscriptomic data we observe that *Ruminococcus bicirculans, R. callidus*, and *R. champallensis* have actually lower expression in T2D patients, and only sp. DSM 100440 has higher prevalence and expression in T2D patients (see Supplementary Table s1 and Supplementary Figure s2a). In the literature, there is a prevalent notion that at the genus level, *Blautia* is positively associated with T2D. In our data, with much higher resolution, we are able to observe that out of the 8 species, *Blautia massiliensis* and *Blautia Marseille-P3087* have lower prevalence and expression in T2D as opposed to the non-T2D while there are six other species that have indeed higher prevalence and expression in Type 2 Diabetes patients (see Supplementary Table s2 and Supplementary Figure s2b).

### Diagnostic Model and Risk Score

To pick up early signals of metabolic disease, we performed statistical analysis to derive features that are associated with all the subjects with T2D vs. the general population who may have other comorbidities and may be subjected to different types of treatments, and separately performed ML analysis to derive a risk score for T2D. As shown in Supplementary Figure S4b, taxa and KO diversity decrease in the T2D patients (KO diversity is significant p=3.27e-07). We present the taxonomic composition using the differential prevalence (based on Chi-square method) (see Supplementary Figure S6c) and KOs (see Supplementary Figure S6e) of the microbiomes of Type 2 Diabetes patients vs. individuals in the general population. In Supplementary Figure S6d) we present the differential expression of taxa and in Supplementary S6f) functional changes using differential expression based on Kruskal-Wallis method. We observe that there is a great overlap between the highest prevalent taxa with the statistical test that we presented in Figure 2, as 65 species are in common (out of 266 species) with the statistically significant species from the general population vs T2D, and yet, there are 201 unique species, which is due to the fact that there is a very big variability across this general population where there are influences from different comorbidities as well as various types of medications taken. Similarly, we observed that 99 (out of 338 species) that are differentially expressed are statistically significant both in the non-medicated vs. T2D as well as in the general population vs. T2D cohorts (taxa and KO overlaps are presented in Supplementary Figure S6g-j). We developed a general classifier with 922 KOs and species which is informative to provide a risk score that can predict whether an individual has a high risk of being diagnosed with T2D using the gut microbiome (see Figure 3a and Supplementary Figure S7a-b). As shown in Figure 3b), in the validation cohort (n=3028) our model can distinguish between type 2 diabetes (mean=62.81) and non-diabetic (mean=44.10) individuals (t-test, p=2.41e-47), as well as between prediabetic(mean=53.25) and non-diabetic (p=2.88e-24) and also between T2D vs. pre-diabetic individuals (p=6.76e-07). We observe a weak correlation between the T2D score and BMI of the validation cohort (Rho=0.16, p=5.29e-19) (see Supplementary Figure S7c), which suggests that the BMI alone is not sufficient and a predictive model is still needed to predict the risk of developing T2D.

**Figure 3.**
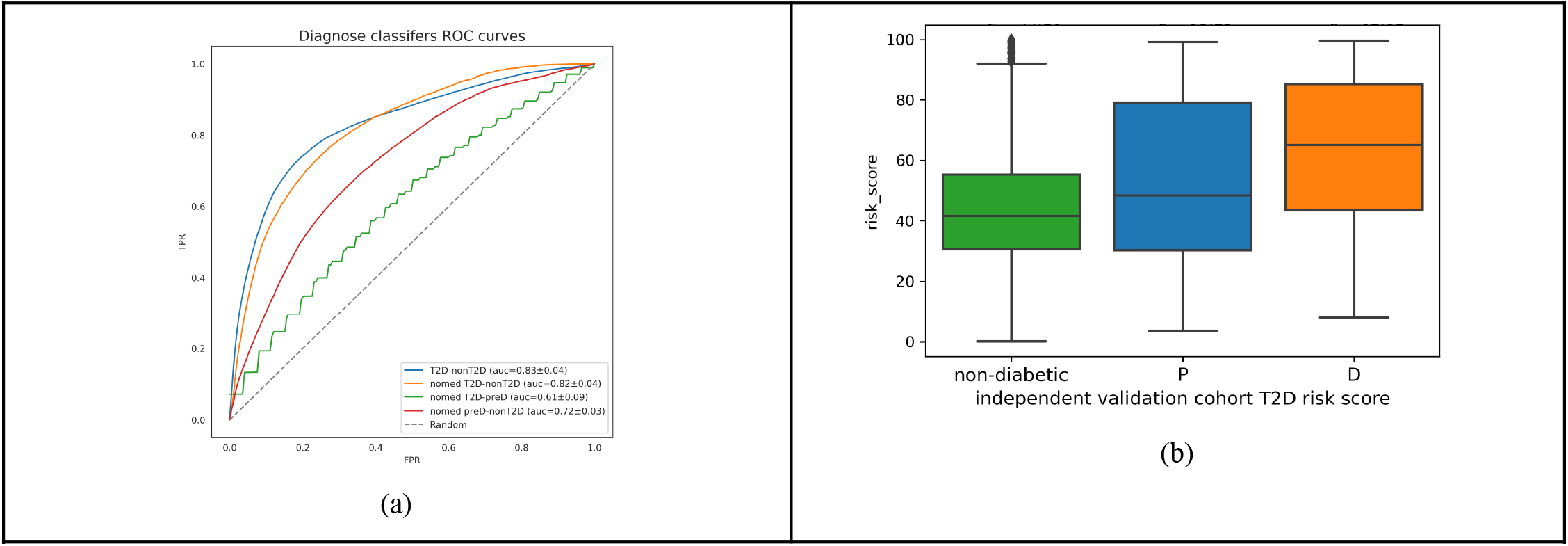
Type 2 diabetes diagnostic analysis and support vector machine approach. a) ROC curves of diagnostic classifiers b) Risk scores of the validation cohort.

### Metformin control of Type 2 Diabetes

In supplementary table S3 we present all the treatment related comparison cohorts and relevant analyses. Due to the limited number of samples we notice that the statistical analysis does not yield results except in the metformin related analysis there are two statistically significant KOs, namely K01273 and K02760 which show higher differential prevalence in non-responders (p<0.05). However, we are able to get strong signals from an SVM approach yielding ROC-AUC 0.74 when we compare patients who are taking metformin and are able to control their HbA1c (<6.5), vs. patients who are unable to control their HbA1c (>=6.5) (see figure 4a).

**Figure 4:**
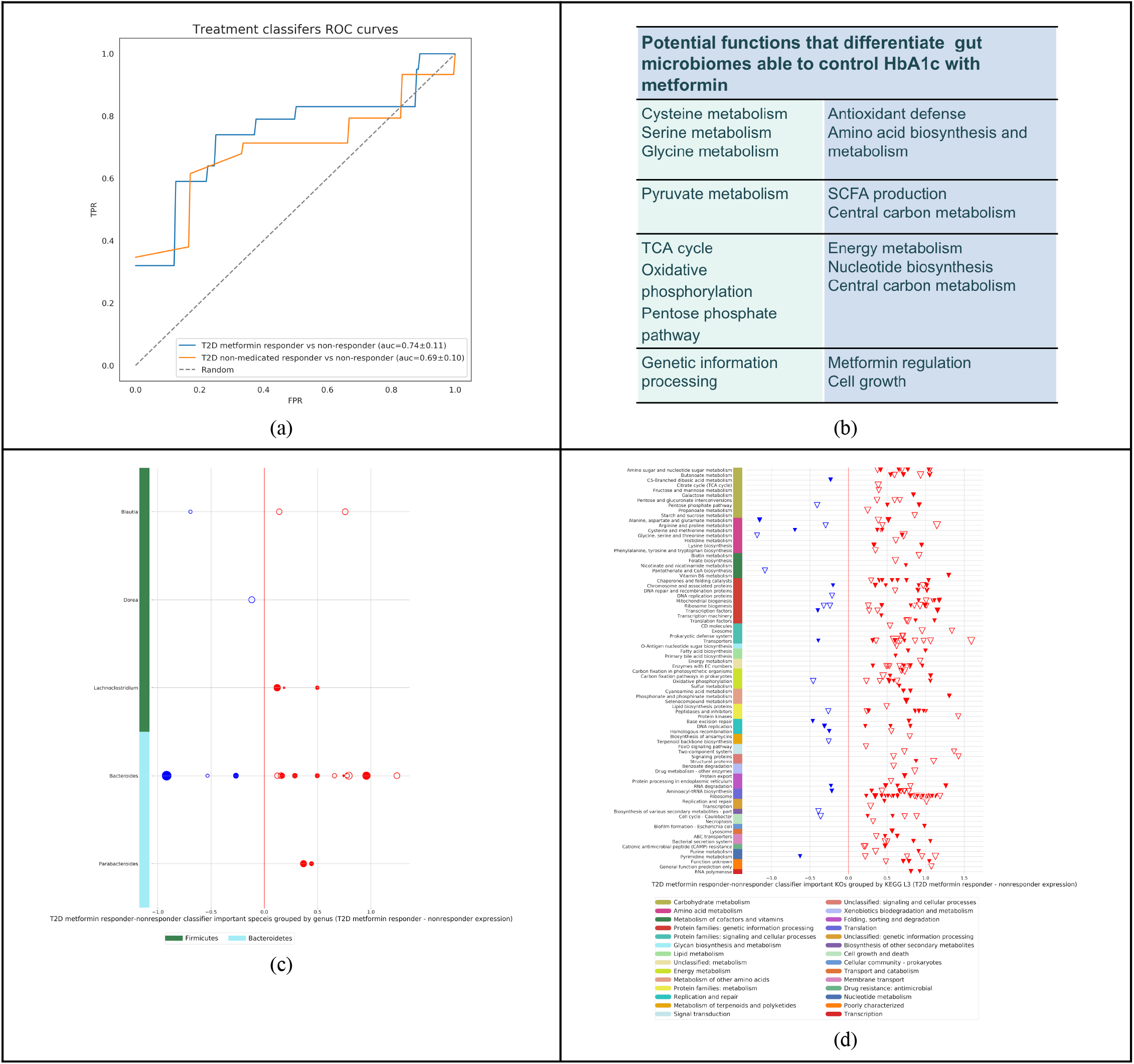
T2D metformin treated patients with good control of HbA1c (<6.5) (n=22) vs T2D metformin treated patients who cannot control their HbA1c (>=6.5) (n=41) AUC = 0.74+/-0.11 (a) ROC curves of metformin treatment classifiers. (b) Summary of important features and potential functions of the metformin models. (c) Important differentially expressed species in T2D non-medicated patients who are able to control HbA1c (<=6.5) in red vs. patients who are non-medicated and not able to control HbA1c (>6.5) in blue. (d) Differentially expressed KOs in T2D non-medicated patients who are able to control HbA1c (<=6.5) in red vs. patients who are non-medicated and not able to control HbA1c (>6.5) in blue. Here we visualize species with differential expression > 0.1; top 300 KOs with the highest importance coefficients and differential expression > 0.2.

Supplementary Figure S8 presents our results in richness (see Supplementary Figure S8a) and diversity (see Supplementary Figure S8b) between Metformin-treated-HBA1c controlled and the non-controlled groups, and we observe higher and statistically significant differences in KO richness and species richness in the non-controlled group. We present the scatter plots in Figure 4.

We also get strong signals when we compare individuals who are not taking any medications and instead rely on diet and lifestyle changes and are able to control their HbA1c (<6.5), vs. patients who are unable to control their HbA1c (>=6.5) (see Figure 4a). Supplementary Figure S8 presents our results in richness (see Supplementary Figure S8c) and diversity (see Supplementary Figure S8d) between non-medicated-HbA1c controlled groups, and we observed higher statistically significant level for species diversity (p=3.06e-02), for patients who are able to control HbA1c. Our SVM approach yielded a classifier showing ROC-AUC 0.69 (see Figure 4a). We present the scatter plots of the features in Supplementary Figure S8.

## Discussion

Literature abounds with metagenomic studies over the past decade, and many genera have been associated with T2D (Gurung et al., 2020). Our study observes certain functions and species related to opportunistic pathogens, lipopolysaccharides (LPS), oxidative stress, and osmotic stress, as associated with T2D. Active signatures associated with the non-diabetic cohort, such as taxa capable of SCFA and succinate production, are also identified (in the Supplementary). We summarize the observations from our data in Figure 5 and in the following section.

**Figure 5.**
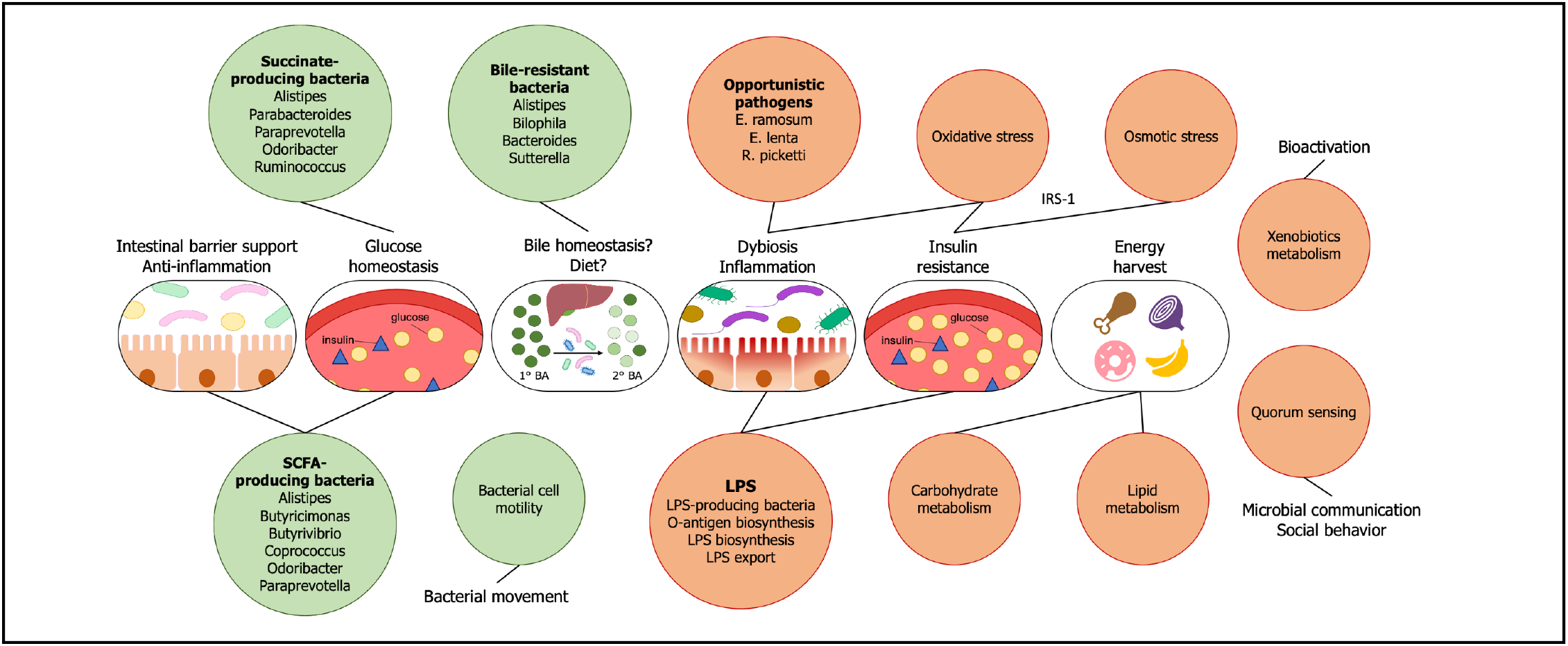
Biological interpretation of the type 2 diabetes metatranscriptomic analysis and derived features. Orange and green circles represent features enriched in the diseased (preT2D or T2D) and non-diabetic cohorts, respectively.

### Taxonomical Shifts in the T2D Metatranscriptome

Studies have demonstrated the alterations of gut microbiota composition in T2D and suggested that gut dysbiosis is a factor in the development of insulin resistance (Allin et al., 2015; Sircana et al., 2018). Our statistical model reveals specific *Firmicutes* and *Proteobacteria* features enriched in both the prediabetes and T2D samples when compared to the presumed healthy population (Figure 2a, S1a, and S1b). Although many of these taxa are uncharacterized, some have been linked to T2D in metagenomics studies while others are known opportunistic pathogens associated with altered metabolic phenotypes. *E. ramosum*, a feature observed in our diseased cohorts, is an opportunistic bacterium that may promote intestinal absorption of glucose and fat in obesity (Mandić et al., 2019; Woting et al., 2014), has also been shown to be T2D-enriched in a metagenomic study of a Chinese cohort (Qin et al., 2012). Higher prevalence and abundance of *Eggerthella* and *Ralstonia* features, including *E. lenta* and *R. pickettii*, are also observed in our data. The opportunistic *E. lenta* has been reported to be enriched in the T2D microbiome, and *R. pickettii* has been suggested to aggravate glucose intolerance in obesity (Qin et al., 2012; Udayappan et al., 2017). Overall, the taxonomical shifts in our diseased cohorts highlight the presence of opportunistic pathogens and their potentials as T2D progression markers. On the other hand, the differential expression of SCFA-producing taxa, succinate-producing taxa, and bile-resistant microorganisms is noted in the non-diabetic cohort and discussed further in the Supplement. A detailed discussion of other T2D-enriched taxa is included in the Supplement.

### Lipopolysaccharide-Associated and Proinflammatory Features

One of the mechanisms through which the gut microbiota interacts with the host is through the production and shedding of LPS, which elicits a pro-inflammatory cascaded response of the immune system. Metabolic endotoxemia, as a result of LPS translocation across the intestinal barrier and into the bloodstream, causes inflammation and is intertwined with the development of insulin resistance (Cani et al., 2007). In our study, this phenomenon is best highlighted by several LPS-producing species identified to be enriched in the T2D cohort, such as *R. picketti* and *Escherichia albertii* (Figure 2a & S1a). The fecal abundance of *R. picketti* has been shown to increase in obese patients with impaired glucose tolerance and type 2 diabetes, and the organism has been suggested to play a role in the development of glucose intolerance and increased inflammatory markers (Udayappan et al., 2017). *E. albertii* is an enteropathogen increasingly associated with diarrhea and gastroenteritis. In addition, the abundance of *Escherichia spp*. has been shown to increase as a result of metformin treatment (Forslund et al., 2015). Functionally, the model also identifies KOs involved in the biosynthesis of O-antigen, the exterior constituent of LPS and often contributing to virulence, and the biosynthesis, regulation, and export of LPS, particularly in the prediabetes population.

### Overall Functional Characteristics

Previous metagenome-based studies report the enrichment of cell motility and flagellar assembly pathways in non-diabetic individuals (Karlsson et al., 2013; Qin et al., 2012). This finding is corroborated by our analysis using metatranscriptomics, with bacterial motility proteins including those involved in chemotaxis and pilus assembly identified to be differentially expressed in the non-diabetic cohort, particularly in the comparison to the T2D cohort. Our T2D samples are functionally enriched with activities from carbohydrate metabolism, glycerolipid metabolism, glycerophospholipid metabolism, transporters, and xenobiotics degradation. In addition, unique features such as quorum sensing and ribosome biogenesis are also differentially expressed in the T2D cohort (Figure 2b).

### Features of Osmotic and Oxidative Stress in Prediabetes and T2D

Functional analyses of T2D metagenomes using Chinese and European cohorts have revealed the enrichment of transporters for sugars, amino acids, and ions (Karlsson et al., 2013; Qin et al., 2012). We find in our sampled population that transporter activities, especially those of ions, are differentially prevalent and expressed in the microbiome of the diseased cohorts compared to that of the non-diabetes. In addition, the high prevalence of osmolyte transporters is observed within both the preT2D and T2D cohorts, although the signal particularly is strong in the former, where several KOs related to glycine betaine transport are identified. Elevated plasma tonicity is thought to be a risk factor for diabetes progression in patients with hyperglycemia (Stookey et al., 2004), while hyperosmotic stress has been shown in adipocytes to suppress insulin action through the serine phosphorylation of insulin receptor substrate-1 (IRS1), leading to cellular insulin resistance (Gual et al., 2003). We therefore surmise that the prediabetic gut microbiome experiences local osmotic stress, and by actively uptaking and accumulating osmoprotective compounds such as glycine betaine in the cells, bacteria adapt to osmotic stress in response to osmolarity changes in the environment (Sleator & Hill, 2002).

The active functions of the preT2D cohort largely mirror that of the T2D microbiome in terms of carbohydrate metabolism, lipid metabolism, and transporters. Additionally, we identify functions associated with oxidative stress, including KOs involved in sequestering oxidative damage and glutathione-dependent redox chemistry, to have higher differential expression in preT2D. Oxidative stress can induce inflammation and is known to play a significant role in the progression of type 2 diabetes (Folli et al., 2011). Metabolic endotoxemia has also been shown to promote oxidative stress, while antibiotic treatment reduces its occurrence (P. D. Cani et al., 2008). Previously, metagenomic studies using Chinese and European cohorts have revealed the enrichment of oxidative stress-related functions in T2D patients (Karlsson et al., 2013; Qin et al., 2012); here, we hypothesize that such stress response is discernible by the prediabetic stage at the metatranscriptomic level.

### Functional Features of the Metformin Treatment Model

Changes in microbial functions are thought to play a role in mediating the beneficial effects of diabetes treatments, and metformin has been associated with increased activities in pathways such as SCFA production, lipopolysaccharide biosynthesis, sphingolipid and fatty acid metabolism, transporters, amino acid biosynthesis and metabolism, and pyruvate metabolism (Forslund et al., 2015; Lee & Ko, 2014; Ma et al., 2018; Wu et al., 2017). We summarize the functional findings from the metformin treatment model in Figure 4b and as follows. The analysis of taxa features is included in the Supplement.

Adding to the observation that amino acid metabolism is altered in the microbiome of metformin-treated patients, our model further identifies features in cysteine and methionine metabolism in predicting control over HbA1c. Many of such features are involved in the metabolic reactions surrounding cysteine, a precursor to glutathione. Glutathione is one of the most critical molecules in the defense against oxidative stress. Low plasma cysteine level and the resultant decreased glutathione synthesis have been associated with inflammation in IBD patients (Sido et al., 1998). Perhaps in line with the importance of cysteine, several KOs surrounding the serine and glycine nodes are also suggested by the model. These amino acids serve as the precursors not only to each other but also to cysteine. Additionally, glycine is tied to increased insulin sensitivity (Adeva-Andany et al., 2018), and earlier studies have shown that glycine degradation activities are enriched in metformin-untreated T2D patients and in prediabetic exercise non-responders (Forslund et al., 2015; Liu et al., 2020). Our treatment model also identifies some features in lysine biosynthesis to be important. The butyrogenic property of lysine has been demonstrated by a human gut commensal (Bui et al., 2015), but more information is required to clarify how microbially produced lysine is linked to T2D.

An earlier metagenomic study has demonstrated the reduced pyruvate synthase capacity in metformin-untreated T2D samples (Forslund et al., 2015). In another study combining fecal metagenomic data from T2D patients and metatranscriptomic data from *in vitro* gut simulators, pyruvate metabolism has been found to be enriched with metformin treatment in both types of analysis (Wu et al., 2017). From our model, KOs surrounding the pyruvate node are identified to be important; along with several features involved in the TCA cycle, the importance of the central carbon metabolism is thus highlighted. Although it is unclear how these features contribute to metformin response, a possible explanation may be the accompanying production of SCFAs or organic acids such as succinate from pyruvate fermentation. The importance of energy metabolism and nucleotide biosynthesis in metformin-responders is also highlighted by multiple KOs from the TCA cycle, oxidative phosphorylation, and pentose phosphate pathway (Figure 4b).

Applications include a stool test that can predict whether metformin will work for a particular individual. Furthermore, the gut microbiome can potentially be modulated to make metformin more effective.

## Conclusions

The richness of the metatranscriptomic data also enabled us to develop and validate a T2D risk model, with a score that can distinguish individuals with glycemic dysregulation (prediabetes and diabetes) from those with normal glucose metabolism.

Our methodologies include clinical grade laboratory analyses and bioinformatics, machine learning, and advanced statistical approaches leveraging data from 53,947 individuals. While some of our findings are consistent with the published literature, we observe novel associations as our metatranscriptome approach allows for more direct functional observations of the human gut microbiome.

Our metatranscriptomic analysis illuminates the significance of both the taxa and microbial pathways pertaining to opportunistic pathogens, lipopolysaccharides (LPS), lipid metabolism, and ion transporters, as well as implications for inflammation, oxidative stress, and osmotic stress in the diseased cohorts. In the non-diabetic, presumed-healthy population, our analysis highlights the elevated activity of features associated with bile resistance, short chain fatty acids (SCFAs) production, cell motility, and succinate production that have not been reported before at the metatranscriptomic level.

Additionally, our machine learning model was able to distinguish novel metatranscriptomic features that segregate patients who receive metformin and are able to control their HbA1c from those who do not. Our model points to complex changes in the amino acid metabolism, pyruvate metabolism, TCA cycle, and oxidative phosphorylation, as well as proteins related to genetic information processing important in determining treatment response.

## Data Availability

Summary data is available within the Article and Supplementary Information. Research requiring additional data can visit https://www.viome.com/vri/data-access, through which we provide access to summary statistics through a Data Transfer Agreement that protects the privacy of our participants' data.

## Author contributions

N.D. conceived the phenotype stratification, guided the analysis based on it, and led the strategy for the manuscript. N.S. performed all the data analyses and visualizations, with support from Y.C. C.H.H. interpreted the features presented in the discussion. M.V. supported with the laboratory analyses; P.J.T. and F.C. supported with the bioinformatic analyses. D.T. provided clinical perspective and validated the results. All others contributed to editing the manuscript. G.B. provided overall direction and did the final editing of the manuscript.

## Acknowledgments

The authors thank Iordan Slavov and Hal Tily for early versions of type 2 diabetes analyses. We also thank Pedro Moura for engineering the system to collect metadata from Viome customers for this study.

## Methods

### Study participant recruitment

The data analyzed for the purpose of this report was obtained from Viome customers, who either completed a research Informed Consent Form (approved by a federally-accredited Institutional Review Board), or agreed to have their data analyzed in the terms and conditions, during the purchase of the gut microbiome test. All study data are de-identified; the laboratory, bioinformatics, and data science team members never had access to any personally identifiable information.

### Sample collection and laboratory analysis

The metatranscriptomic method that we use is designed for large-scale population analysis of stool samples as described previously (Hatch et al., 2019), and included sample collection, ambient temperature sample preservation, total RNA extraction, physical removal of ribosomal RNAs (rRNAs), preparation of directional Illumina libraries, and Illumina sequencing. The stability of the RNA stabilizer was tested for up to 28 days at ambient temperature, including shipping.

### Phenotype data collection

The clinical phenotypes and medication status were labeled based on the answers to the Viome questionnaire. The non-diabetic group includes presumed healthy individuals who are not reporting medical conditions as well as individuals who have reported comorbidities, while excluding anyone taking antibiotics within one month, proton pump inhibitor and/or acid suppressants, had abdominal surgery or with diseases that may affect gut microbiome (e.g. IBS, IBD, colon cancer). For this group we have matched the age, sex and BMI to those of prediabetic and T2D group. The first stratification is based on the type of diagnosis that patients report, including pre-diabetes and T2D. Within the prediabetic and T2D groups there are patients who receive treatment in the form of medications, as well as patients who are not on medications. We consider within the T2D group that subjects who have reported HbA1c <6.5 to be responding to the treatment, and the patients with >=6.5 to be the non-responders.

### Bioinformatics processing

Paired-end reads were mapped to a catalog of 53,660 microbial genome assemblies spanning archaea, bacteria, fungi, protozoa, and viruses. (We downloaded the complete genomes available in NCBI Reference Sequence Database, and used the GenBank sequence database for viral genomes.) Strain-level relative activities were computed from mapped reads via the expectation-maximization (EM) algorithm (Dempster et al., 1977). Relative activities at other levels of the taxonomic tree were then computed by aggregation according to the taxonomic rank. Relative activities for biological functions were computed by mapping paired-end reads to a catalog of 52,324,420 microbial genes, quantifying gene-level relative activities with the EM algorithm, and then aggregating gene-level activity by KEGG Ortholog (KO) annotation (Kanehisa & Goto, 2000). The identified and quantified active microbial species and KOs for each sample were then provided to the T2D classifier. (More details in Supplementary Material.)

### Descriptive statistical analysis

Standard statistical analyses described below were initially performed to analyze the differential expression of active microorganisms and active functions between. The data was transformed using the centered log ratio transformation (CLR) (Aitchison, 1982) after imputation of zero values using multiplicative replacement (Martín-Fernández et al., 2003). The age, sex and BMI of non-diabetic controls were matched to those of samples with T2D or pre-diabetics. We used the Mann-Whitney U (MWU) test (FDR < 0.05) to evaluate differential expression and Chi-2 test (FDR < 0.05) to evaluate differential prevalence of species or KOs between different groups. It is important to note that this is a descriptive statistical test to analyze features independently for differential expression without taking into account the interactions among features, and is thus not suitable for the machine learning classification method (below).

### Mapping KOs to functional categories for presentation

For KO visualization scatterplots, the Python module “Bio.KEGG” was used to take as input the KO name and return KO hierarchy at three different levels (level-2 corresponds to level B, level-3 corresponds to level C, and level-4 corresponds to level D in KEGG). Level 2 and level 3 annotation were assigned using EM algorithms (Dempster et al., 1977) with KOs weighted by absolute values coefficients or reversed p values in log scale.

### Richness and diversity

Richness was estimated as the number of present species or KOs in each sample. Shannon-diversity was computed to evaluate the alpha diversity of samples. All the comparisons between richness or Shannon-diversity were done with independent t-test.

### Machine learning

The microbiome data was transformed using the centered log ratio transformation (CLR) (Aitchison, 1982) after imputation of zero values using multiplicative replacement (Martín-Fernández et al., 2003). Machine-learned models using stool microbiome are Support Vector Machines. We have also tried other models such as logistic regression and random forest, and adopted the model giving the highest AUC. Hyperparameter optimization was done using a nested 5-fold cross-validation on the discovery cohort. The inner layer was used for feature selection, training and tuning. We selected the top 20% features with the highest KW test effect size. The outer layer was used for estimation of model performance. The hyperparameters were scored based on balanced accuracy and the final models were evaluated based on balanced accuracy and AUC. In the diagnostic classifiers, the non-diabetic controls were subsampled with matched age, sex and BMI to samples with T2D or pre-diabetics. The number of subsampled non-diabetic controls was three times that of samples with conditions. For pre-diabetic v.s. T2D classifier and all the treatment classifiers, we did not match the two classes because of the small sample size.

The machine learning models will take in multiple features at the same time. In the SVM model’s view, the contribution of each feature is more complex than that of descriptive analysis which takes one feature at a time. In our results, some models will pick more features enriched in non-diabetic samples. It does not necessarily mean that the model’s only predicting non-diabetic samples. Instead, these features are enriched in non-diabetic samples and were also features with lower expression in diabetic samples.

## References

https://doi.org/10.1016/S0140-6736(16)00618-8

Adeva-Andany, M., Souto-Adeva, G., Ameneiros-Rodríguez, E., Fernández-Fernández, C., Donapetry-García, C., & Domínguez-Montero, A. (2018). Insulin resistance and glycine metabolism in humans. Amino Acids, 50(1), 11–27. https://doi.org/10.1007/s00726-017-2508-0

Aitchison, J. (1982). The Statistical Analysis of Compositional Data. Journal of the Royal Statistical Society: Series B (Methodological), 44(2), 139–160. https://doi.org/10.1111/j.2517-6161.1982.tb01195.x

Allin, K. H., Nielsen, T., & Pedersen, O. (2015). Mechanisms in endocrinology: Gut microbiota in patients with type 2 diabetes mellitus. European Journal of Endocrinology, 172(4), R167–177. https://doi.org/10.1530/EJE-14-0874

Bailes, B. K. (2002). Diabetes mellitus and its chronic complications. AORN Journal, 76(2), 266–276, 278–282; quiz 283–286. https://doi.org/10.1016/s0001-2092(06)61065-x

Bui, T. P. N., Ritari, J., Boeren, S., de Waard, P., Plugge, C. M., & de Vos, W. M. (2015). Production of butyrate from lysine and the Amadori product fructoselysine by a human gut commensal. Nature Communications, 6(1), 10062. https://doi.org/10.1038/ncomms10062

Cani, P. D., Bibiloni, R., Knauf, C., Waget, A., Neyrinck, A. M., Delzenne, N. M., & Burcelin, R. (2008). Changes in Gut Microbiota Control Metabolic Endotoxemia-Induced Inflammation in High-Fat Diet-Induced Obesity and Diabetes in Mice. Diabetes, 57(6), 1470–1481. https://doi.org/10.2337/db07-1403

Cani, Patrice D., Amar, J., Iglesias, M. A., Poggi, M., Knauf, C., Bastelica, D., Neyrinck, A. M., Fava, F., Tuohy, K. M., Chabo, C., Waget, A., Delmée, E., Cousin, B., Sulpice, T., Chamontin, B., Ferrières, J., Tanti, J.-F., Gibson, G. R., Casteilla, L., … Burcelin, R. (2007). Metabolic Endotoxemia Initiates Obesity and Insulin Resistance. Diabetes, 56(7), 1761–1772. https://doi.org/10.2337/db06-1491

Chatterjee, S., Khunti, K., & Davies, M. J. (2017). Type 2 diabetes. Lancet (London, England), 389(10085), 2239–2251. https://doi.org/10.1016/S0140-6736(17)30058-2

Dempster, A. P., Laird, N. M., & Rubin, D. B. (1977). Maximum Likelihood from Incomplete Data Via the EM Algorithm. Journal of the Royal Statistical Society: Series B (Methodological), 39(1), 1–22. https://doi.org/10.1111/j.2517-6161.1977.tb01600.x

Folli, F., Corradi, D., Fanti, P., Davalli, A., Paez, A., Giaccari, A., Perego, C., & Muscogiuri, G. (2011). The role of oxidative stress in the pathogenesis of type 2 diabetes mellitus micro- and macrovascular complications: Avenues for a mechanistic-based therapeutic approach. Current Diabetes Reviews, 7(5), 313–324. https://doi.org/10.2174/157339911797415585

Forslund, K., Hildebrand, F., Nielsen, T., Falony, G., Le Chatelier, E., Sunagawa, S., Prifti, E., Vieira-Silva, S., Gudmundsdottir, V., Pedersen, H. K., Arumugam, M., Kristiansen, K., Voigt, A. Y., Vestergaard, H., Hercog, R., Costea, P. I., Kultima, J. R., Li, J., Jørgensen, T., … Pedersen, O. (2015). Disentangling type 2 diabetes and metformin treatment signatures in the human gut microbiota. Nature, 528(7581), 262–266. https://doi.org/10.1038/nature15766

Gual, P., Gonzalez, T., Grémeaux, T., Barres, R., Le Marchand-Brustel, Y., & Tanti, J.-F. (2003). Hyperosmotic stress inhibits insulin receptor substrate-1 function by distinct mechanisms in 3T3-L1 adipocytes. The Journal of Biological Chemistry, 278(29), 26550–26557. https://doi.org/10.1074/jbc.M212273200

Gurung, M., Li, Z., You, H., Rodrigues, R., Jump, D. B., Morgun, A., & Shulzhenko, N. (2020). Role of gut microbiota in type 2 diabetes pathophysiology. EBioMedicine, 51, 102590. https://doi.org/10.1016/j.ebiom.2019.11.051

Hatch, A., Horne, J., Toma, R., Twibell, B. L., Somerville, K. M., Pelle, B., Canfield, K. P., Genkin, M., Banavar, G., Perlina, A., Messier, H., Klitgord, N., & Vuyisich, M. (2019). A Robust Metatranscriptomic Technology for Population-Scale Studies of Diet, Gut Microbiome, and Human Health. International Journal of Genomics, 2019. https://doi.org/10.1155/2019/1718741

Hu, F. B. (2011). Globalization of Diabetes: The role of diet, lifestyle, and genes. Diabetes Care, 34(6), 1249–1257. https://doi.org/10.2337/dc11-0442

Kanehisa, M., & Goto, S. (2000). KEGG: Kyoto encyclopedia of genes and genomes. Nucleic Acids Research, 28(1), 27–30. https://doi.org/10.1093/nar/28.1.27

Karlsson, F. H., Tremaroli, V., Nookaew, I., Bergström, G., Behre, C. J., Fagerberg, B., Nielsen, J., & Bäckhed, F. (2013). Gut metagenome in European women with normal, impaired and diabetic glucose control. Nature, 498(7452), 99–103. https://doi.org/10.1038/nature12198

Larsen, N., Vogensen, F. K., van den Berg, F. W. J., Nielsen, D. S., Andreasen, A. S., Pedersen, B. K., Al-Soud, W. A., Sørensen, S. J., Hansen, L. H., & Jakobsen, M. (2010). Gut microbiota in human adults with type 2 diabetes differs from non-diabetic adults. PloS One, 5(2), e9085. https://doi.org/10.1371/journal.pone.0009085

Lawlor, N., George, J., Bolisetty, M., Kursawe, R., Sun, L., Sivakamasundari, V., Kycia, I., Robson, P., & Stitzel, M. L. (2017). Single-cell transcriptomes identify human islet cell signatures and reveal cell-type-specific expression changes in type 2 diabetes. Genome Research, 27(2), 208–222. https://doi.org/10.1101/gr.212720.116

Lee, H., & Ko, G. (2014). Effect of metformin on metabolic improvement and gut microbiota. Applied and Environmental Microbiology, 80(19), 5935–5943. https://doi.org/10.1128/AEM.01357-14

Ley, R. E., Turnbaugh, P. J., Klein, S., & Gordon, J. I. (2006). Microbial ecology: Human gut microbes associated with obesity. Nature, 444(7122), 1022–1023. https://doi.org/10.1038/4441022a

Li, Q., Chang, Y., Zhang, K., Chen, H., Tao, S., & Zhang, Z. (2020). Implication of the gut microbiome composition of type 2 diabetic patients from northern China. Scientific Reports, 10(1), 5450. https://doi.org/10.1038/s41598-020-62224-3

Liu, Y., Wang, Y., Ni, Y., Cheung, C. K. Y., Lam, K. S. L., Wang, Y., Xia, Z., Ye, D., Guo, J., Tse, M. A., Panagiotou, G., & Xu, A. (2020). Gut Microbiome Fermentation Determines the Efficacy of Exercise for Diabetes Prevention. Cell Metabolism, 31(1), 77-91.e5. https://doi.org/10.1016/j.cmet.2019.11.001

Ma, W., Chen, J., Meng, Y., Yang, J., Cui, Q., & Zhou, Y. (2018). Metformin Alters Gut Microbiota of Healthy Mice: Implication for Its Potential Role in Gut Microbiota Homeostasis. Frontiers in Microbiology, 9, 1336. https://doi.org/10.3389/fmicb.2018.01336

Mandić, A. D., Woting, A., Jaenicke, T., Sander, A., Sabrowski, W., Rolle-Kampcyk, U., von Bergen, M., & Blaut, M. (2019). Clostridium ramosum regulates enterochromaffin cell development and serotonin release. Scientific Reports, 9(1), 1177. https://doi.org/10.1038/s41598-018-38018-z

Martín-Fernández, J. A., Barceló-Vidal, C., & Pawlowsky-Glahn, V. (2003). Dealing with Zeros and Missing Values in Compositional Data Sets Using Nonparametric Imputation. Mathematical Geology, 35(3), 253–278. https://doi.org/10.1023/A:1023866030544

Qin, J., Li, Y., Cai, Z., Li, S., Zhu, J., Zhang, F., Liang, S., Zhang, W., Guan, Y., Shen, D., Peng, Y., Zhang, D., Jie, Z., Wu, W., Qin, Y., Xue, W., Li, J., Han, L., Lu, D., … Wang, J. (2012). A metagenome-wide association study of gut microbiota in type 2 diabetes. Nature, 490(7418), 55–60. https://doi.org/10.1038/nature11450

Ridaura, V. K., Faith, J. J., Rey, F. E., Cheng, J., Duncan, A. E., Kau, A. L., Griffin, N. W., Lombard, V., Henrissat, B., Bain, J. R., Muehlbauer, M. J., Ilkayeva, O., Semenkovich, C. F., Funai, K., Hayashi, D. K., Lyle, B. J., Martini, M. C., Ursell, L. K., Clemente, J. C., … Gordon, J. I. (2013). Gut microbiota from twins discordant for obesity modulate metabolism in mice. Science (New York, N.Y.), 341(6150), 1241214. https://doi.org/10.1126/science.1241214

Segerstolpe, Å., Palasantza, A., Eliasson, P., Andersson, E.-M., Andréasson, A.-C., Sun, X., Picelli, S., Sabirsh, A., Clausen, M., Bjursell, M. K., Smith, D. M., Kasper, M., Ämmälä, C., & Sandberg, R. (2016). Single-Cell Transcriptome Profiling of Human Pancreatic Islets in Health and Type 2 Diabetes. Cell Metabolism, 24(4), 593–607. https://doi.org/10.1016/j.cmet.2016.08.020

Sengupta, U., Ukil, S., Dimitrova, N., & Agrawal, S. (2009). Expression-based network biology identifies alteration in key regulatory pathways of type 2 diabetes and associated risk/complications. PloS One, 4(12), e8100. https://doi.org/10.1371/journal.pone.0008100

Sido, B., Hack, V., Hochlehnert, A., Lipps, H., Herfarth, C., & Dröge, W. (1998). Impairment of intestinal glutathione synthesis in patients with inflammatory bowel disease. Gut, 42(4), 485–492. https://doi.org/10.1136/gut.42.4.485

Sircana, A., Framarin, L., Leone, N., Berrutti, M., Castellino, F., Parente, R., De Michieli, F., Paschetta, E., & Musso, G. (2018). Altered Gut Microbiota in Type 2 Diabetes: Just a Coincidence? Current Diabetes Reports, 18(10), 98. https://doi.org/10.1007/s11892-018-1057-6

Sleator, R. D., & Hill, C. (2002). Bacterial osmoadaptation: The role of osmolytes in bacterial stress and virulence. FEMS Microbiology Reviews, 26(1), 49–71. https://doi.org/10.1111/j.1574-6976.2002.tb00598.x

Stookey, J. D., Pieper, C. F., & Cohen, H. J. (2004). Hypertonic hyperglycemia progresses to diabetes faster than normotonic hyperglycemia. European Journal of Epidemiology, 19(10), 935–944. https://doi.org/10.1007/s10654-004-5729-y

Turnbaugh, P. J., Hamady, M., Yatsunenko, T., Cantarel, B. L., Duncan, A., Ley, R. E., Sogin, M. L., Jones, W. J., Roe, B. A., Affourtit, J. P., Egholm, M., Henrissat, B., Heath, A. C., Knight, R., & Gordon, J. I. (2009). A core gut microbiome in obese and lean twins. Nature, 457(7228), 480–484. https://doi.org/10.1038/nature07540

Udayappan, S. D., Kovatcheva-Datchary, P., Bakker, G. J., Havik, S. R., Herrema, H., Cani, P. D., Bouter, K. E., Belzer, C., Witjes, J. J., Vrieze, A., de Sonnaville, E. S. V., Chaplin, A., van Raalte, D. H., Aalvink, S., Dallinga-Thie, G. M., Heilig, H. G. H. J., Bergström, G., van der Meij, S., van Wagensveld, B. A., … Nieuwdorp, M. (2017). Intestinal Ralstonia pickettii augments glucose intolerance in obesity. PloS One, 12(11), e0181693. https://doi.org/10.1371/journal.pone.0181693

Wang, H., Lu, Y., Yan, Y., Tian, S., Zheng, D., Leng, D., Wang, C., Jiao, J., Wang, Z., & Bai, Y. (2020). Promising Treatment for Type 2 Diabetes: Fecal Microbiota Transplantation Reverses Insulin Resistance and Impaired Islets. Frontiers in Cellular and Infection Microbiology, 9. https://doi.org/10.3389/fcimb.2019.00455

Wellen, K. E., & Hotamisligil, G. S. (2005). Inflammation, stress, and diabetes. The Journal of Clinical Investigation, 115(5), 1111–1119. https://doi.org/10.1172/JCI25102

Woting, A., Pfeiffer, N., Loh, G., Klaus, S., & Blaut, M. (2014). Clostridium ramosum promotes high-fat diet-induced obesity in gnotobiotic mouse models. MBio, 5(5), e01530–01514. https://doi.org/10.1128/mBio.01530-14

Wu, H., Esteve, E., Tremaroli, V., Khan, M. T., Caesar, R., MannerÅs-Holm, L., StÅhlman, M., Olsson, L. M., Serino, M., Planas-Fèlix, M., Xifra, G., Mercader, J. M., Torrents, D., Burcelin, R., Ricart, W., Perkins, R., Fernàndez-Real, J. M., & Bäckhed, F. (2017). Metformin alters the gut microbiome of individuals with treatment-naive type 2 diabetes, contributing to the therapeutic effects of the drug. Nature Medicine, 23(7), 850–858. https://doi.org/10.1038/nm.4345

Zhang, X., Shen, D., Fang, Z., Jie, Z., Qiu, X., Zhang, C., Chen, Y., & Ji, L. (2013). Human gut microbiota changes reveal the progression of glucose intolerance. PloS One, 8(8), e71108. https://doi.org/10.1371/journal.pone.0071108

Zhou, B., Lu, Y., Hajifathalian, K., Bentham, J., Cesare, M. D., Danaei, G., Bixby, H., Cowan, M. J., Ali, M. K., Taddei, C., Lo, W. C., Reis-Santos, B., Stevens, G. A., Riley, L. M., Miranda, J. J., Bjerregaard, P., Rivera, J. A., Fouad, H. M., Ma, G., … Cisneros, J. Z. (2016). Worldwide trends in diabetes since 1980: A pooled analysis of 751 population-based studies with 4·4 million participants. The Lancet, 387(10027), 1513–1530. https://doi.org/10.1016/S0140-6736(16)00618-8

