## Supplementary materials for "Gut microbiome activity predicts risk of type 2 diabetes and metformin control in a large human cohort"

Figure S1: Progression of disease: statistically important features in a color-bar scatter plot (KW statistics, Chi-square test, FDR p-value < 0.05). Panels (a)-(d) present scatter plots of resulting patient features which include species represented with circles and  KOs represented with triangles.

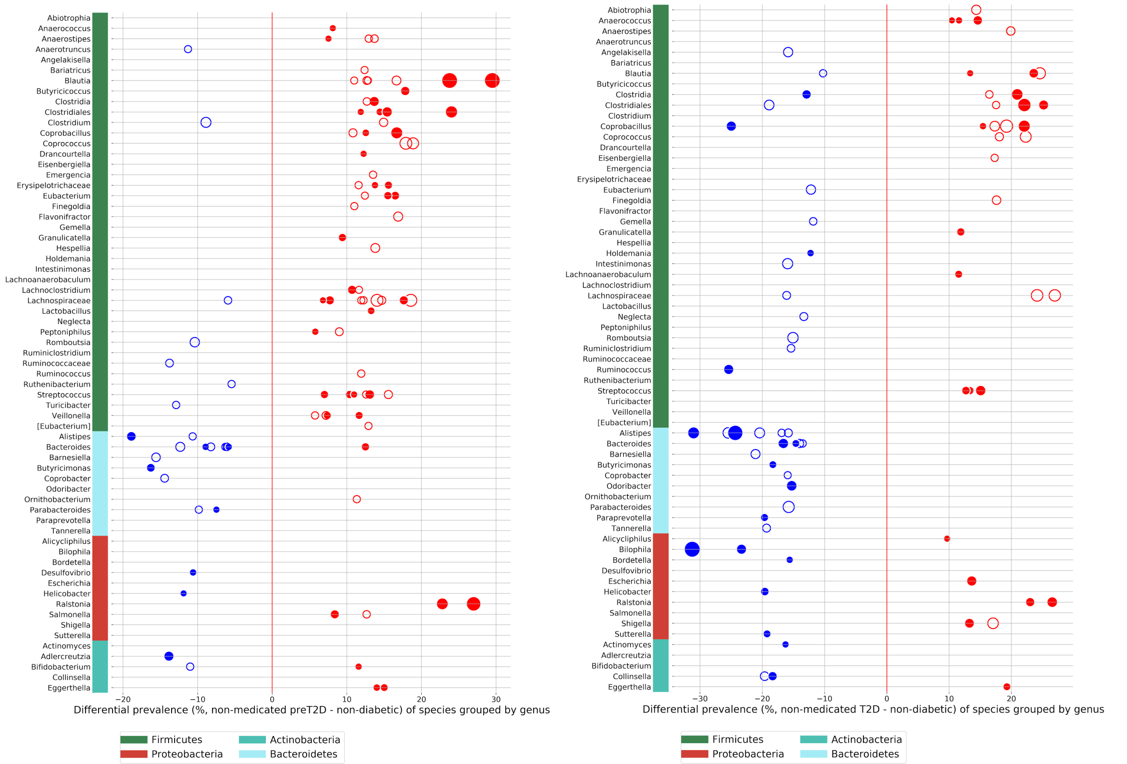


(a) Differential prevalence of species: left panel represents the differential prevalence between pre-T2D vs non-T2D, and the right panel represents the differential prevalence between T2D vs non-T2D

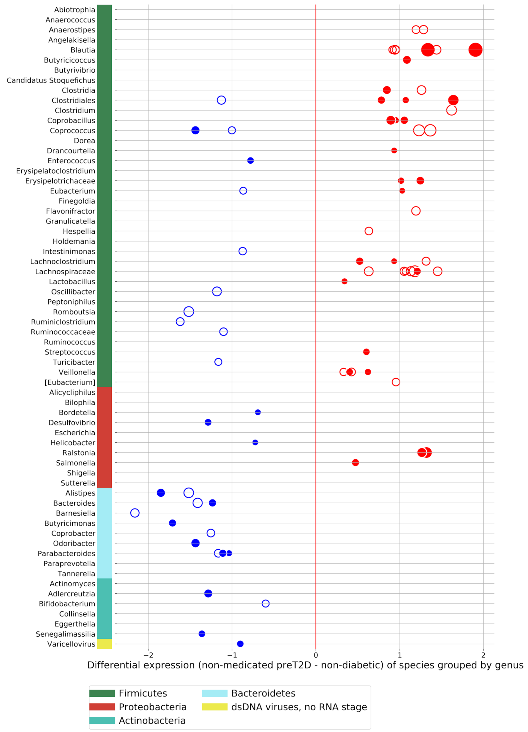


(b) Differential expression of species between pre-T2D vs non-T2D

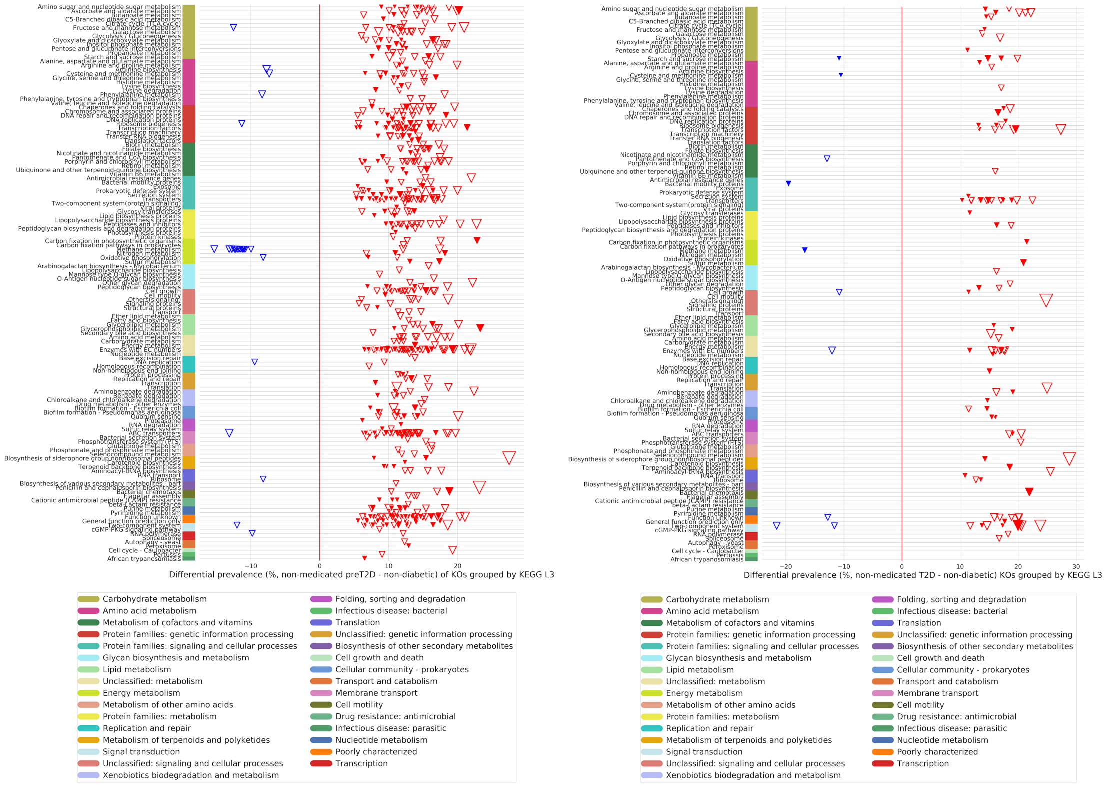


(c) Differential prevalence of KOs, grouped by KEGG category: left panel represents the differential prevalence between non-medicated pre-T2D vs non-medicated non-T2D, and the right side represents the differential prevalence between non-medicated T2D vs non-medicated non-T2D

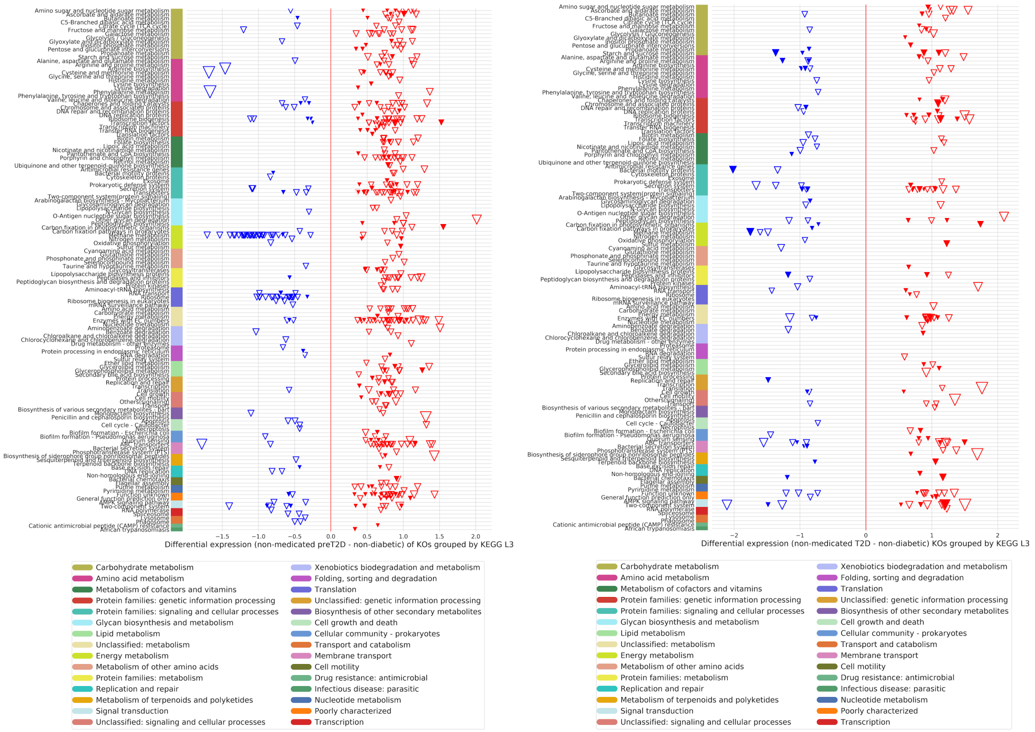


(d) Differential expression of KOs, grouped by KEGG category: left panel represents the differential expression between non-medicated pre-T2D vs non-medicated non-T2D, and the right side represents the differential expression between non-medicated T2D vs non-medicated non-T2D

Table S1. Prevalence and expression of the Ruminococcus species along stages of disease progression.

| Method | Stage | **Ruminococcus bicirculans** | **Ruminococcus callidus** | **Ruminococcus champanellensis** | **Ruminococcus sp. DSM 100440** |
|---|---|---|---|---|---|
| **Prevalence (%)** | **ND** | 68.26 | 57.25 | 39.51 | 30.59 |
| | **Pre-D** | 57.86 | 48.91 | 37.77 | 35.15 |
| | **T2D** | 41.26 | 33.01 | 25.24 | 36.89 |
| **Expression (CLR)** | **ND** | 6.91 +/- 5.58 | 3.79 +/- 4.18 | 1.69 +/- 2.88 | 1.11 +/- 3.08 |
| | **Pre-D** | 5.99 +/- 5.78 | 2.77 +/- 3.81 | 1.55 +/- 2.61 | 1.92 +/- 3.35 |
| | **T2D** | 4.14 +/- 5.73 | 1.90 +/- 3.66 | 0.63 +/- 2.11 | 2.15 +/- 3.72 |

Table S2.Prevalence and expression of the Blautia species along stages of disease progression.

| Method | Stage | **Blautia massiliensis** | **Blautia sp. Marseille-P3087** | **[Ruminococcus] gnavus** | **Blautia sp. Marseille-P3201T** | **Blautia sp. N6H1-15** | **Blautia producta** | **Blautia hansenii** | **Blautia sp. YL58** |
|---|---|---|---|---|---|---|---|---|---|
| **Prevalence (%)** | **ND** | 90.60 | 90.06 | 77.87 | 50.73 | 41.44 | 43.25 | 24.02 | 21.65 |
| | **Pre-D** | 87.34 | 87.99 | 84.28 | 60.48 | 55.68 | 58.73 | 38.21 | 32.97 |
| | **D** | 81.55 | 83.01 | 82.52 | 58.25 | 53.40 | 64.08 | 44.66 | 36.41 |
| **Expression (CLR)** | **ND** | 7.14 +/- 2.75 | 6.73 +/- 2.83 | 5.40 +/- 3.89 | 2.16 +/- 2.93 | 1.65 +/- 3.02 | 1.65 +/- 2.95 | 0.73 +/- 2.49 | 0.39 +/- 2.12 |
| | **Pre-D** | 6.95 +/- 3.06 | 6.54 +/- 2.97 | 6.86 +/- 3.71 | 3.07 +/- 3.09 | 2.62 +/- 3.24 | 3.65 +/- 3.08 | 1.65 +/- 3.30 | 1.77 +/- 2.90 |
| | **D** | 6.17 +/- 3.48 | 5.71 +/- 3.24 | 7.62 +/- 3.91 | 3.17 +/- 3.45 | 2.86 +/- 3.50 | 3.09 +/- 3.07 | 2.52 +/- 3.55 | 1.14 +/- 2.59 |

Figure S2. Bar plots and probability density distribution plots of Ruminococcus and Blautia features

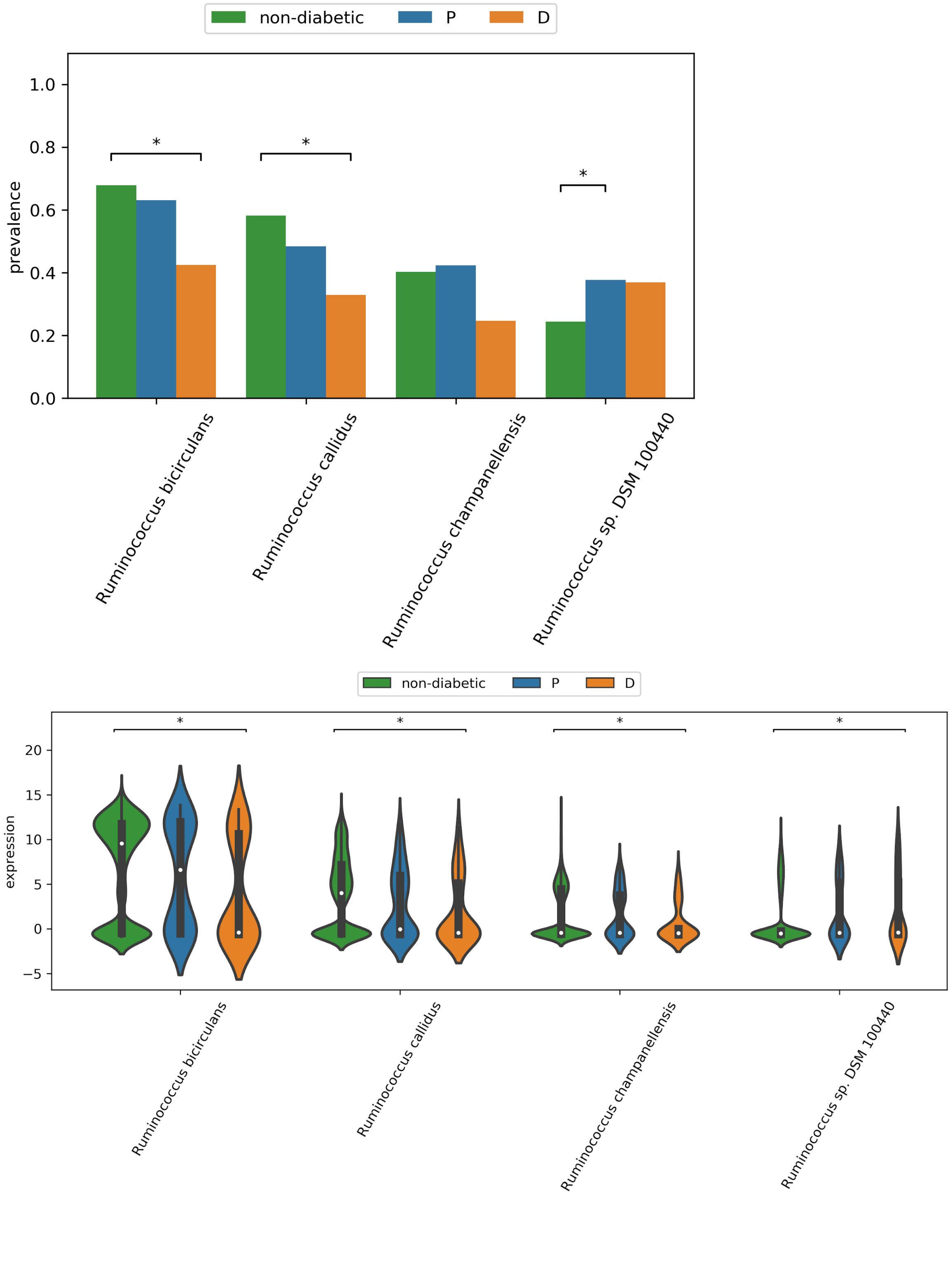


(a) Prevalence bar plot (top) and expression violin plots (bottom) of the species in the Ruminococcus genus

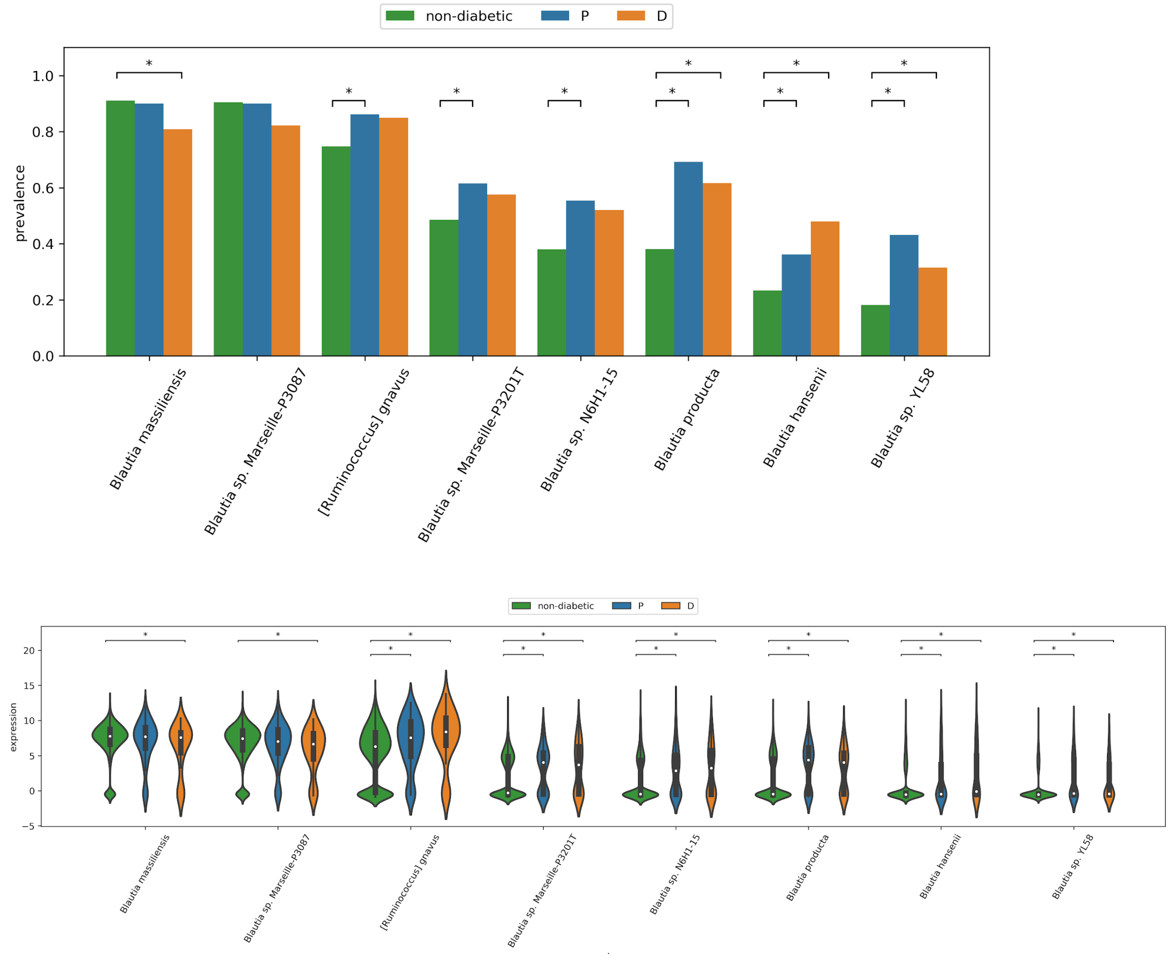


(b) Prevalence bar plot (top) and expression violin plots (bottom) of the species in the Blautia genus

Table S3. Summary of approaches and results for treatment-related cohorts for T2D

| **Comparison** | **Matching criteria** | **Diff. prev. sig. feats.** (p.adj<0.05) | **Diff. exp. sig. feats.** (p.adj<0.05) | **Classification method** | **Mean ROC-AUC** | **Mean balanced accuracy** | **Classifier #features (KOs + species)** |
|---|---|---|---|---|---|---|---|
| T2D metformin HbA1c controlled (n=22) vs T2D metformin HbA1c non-controlled (n=41) | None | 2 KOs 0 species Chi2 test | 0 KOs 0 species KW test | SVM + effect size feature selection | 0.74 | 0.74 | 406 |
| T2D metformin (n=80) vs T2D other drugs (n=23) | None | 0 KOs 0 species Chi2 test | 0 KOs 0 species KW test | SVM + effect size feature selection | 0.56 | 0.52 | 444 |
| T2D metformin (n=80) vs T2D non-medicated (n=79) | None | 0 KOs 0 species Chi2 test | 0 KOs 0 species KW test | SVM + effect size feature selection | 0.62 | 0.61 | 202 |
| T2D non-med "responders" (n=29) vs T2D non-med "non-responder" (n=30) | None | 0 KOs 0 species Chi2 test | 0 KOs 0 species KW test | SVM + effect size feature selection | 0.69 | 0.66 | 408pre-diabetes |
| T2D medicated (n=150) vs T2D non-medicated (n=79) | None | 0 KOs 0 species Chi2 test | 0 KOs 0 species KW test | SVM + effect size feature selection | 0.60 | 0.59 | 449 |
| Pre-T2D medicated (n=34) vs Pre-T2D non-medicated (n=200) | None | 0 KOs 0 species Chi2 test | 0 KOs 0 species KW test | SVM + effect size feature selection | 0.40 | 0.58 | 454 |
| Pre-T2D non-med "responders" (n=63) vs pre-T2D non-med "non-resp" (n=137) | None | 0 KOs 0 species Chi2 test | 0 KOs 0 species KW test | SVM + effect size feature selection | 0.56 | 0.52 | 455 |

Figure S3. Venn diagrams showing intersection of descriptive analysis results of disease progression. Left: comparison of significant differentially prevalent species between preT2D vs. non-diabetic (presumed healthy) in blue, and T2D vs. non-diabetic (presumed healthy) in orange. Right: comparison of significant differentially expressed species between preT2D vs. non-diabetic (presumed healthy) in blue, and T2D vs. non-diabetic (presumed healthy) in orange.

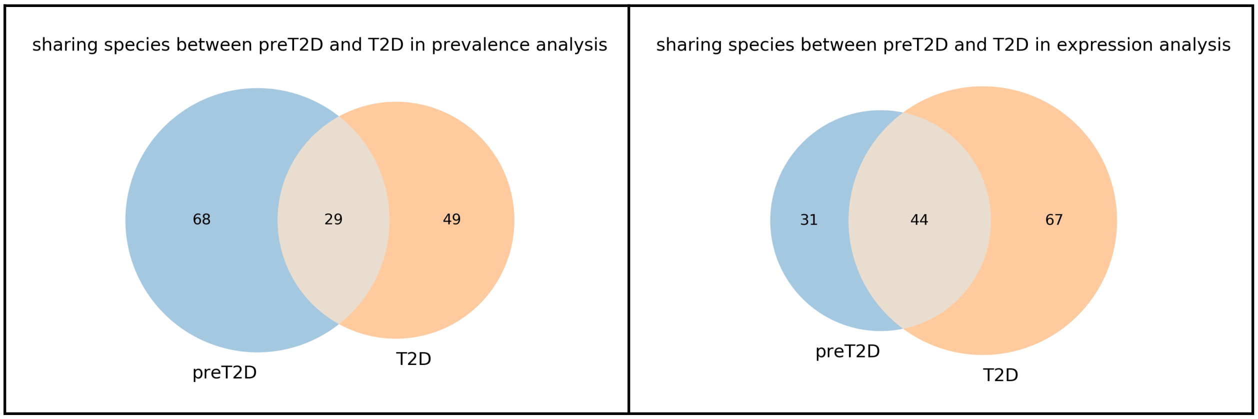


Figure S4. Progression of disease: non-diabetic, pre-daibetic and T2D diversity and richness

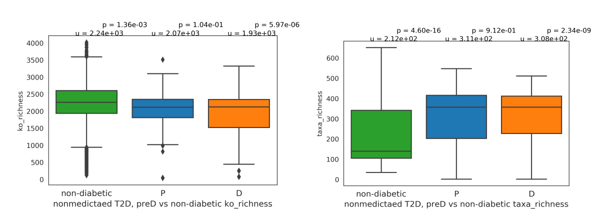


(a) Taxa and KO richness

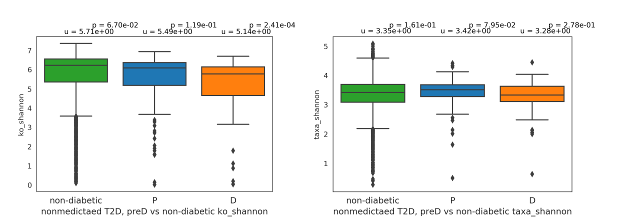


(b) Taxa and KO Shannon diversity

Figure S5. Important features in T2D disease progression classification model distinguishing T2D patients and prediabetic patients from presumed healthy using SVM (species with differential expression > 0.1; top 300 KOs with the highest coefficients and differential expression > 0.2)

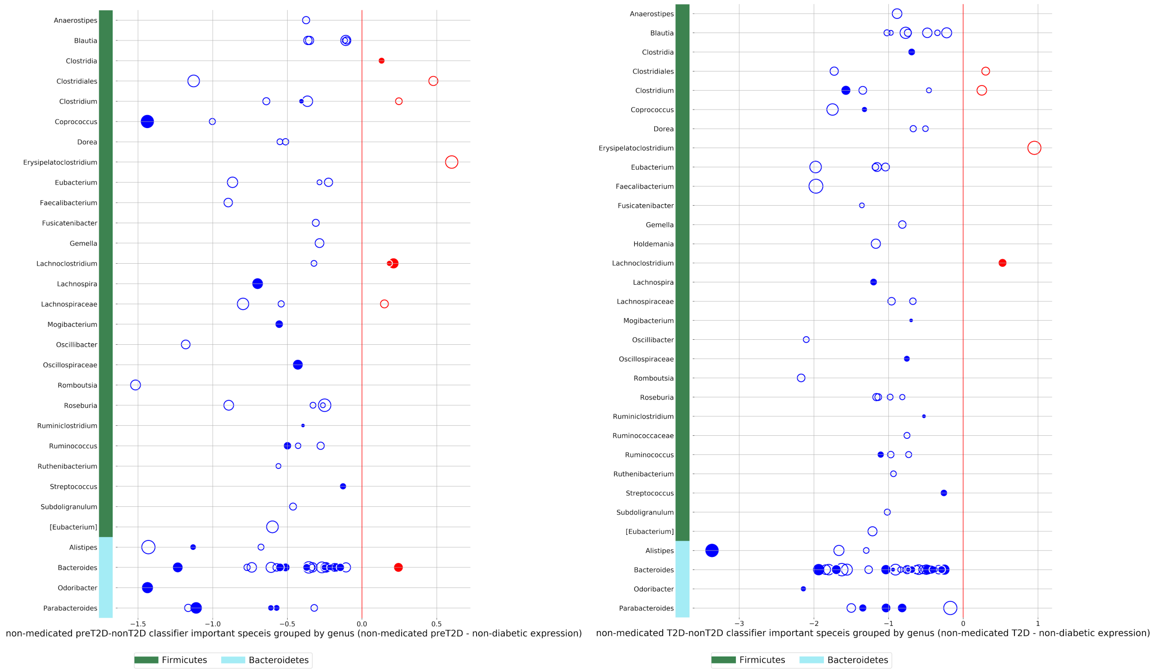


(a) Important species in T2D classifiers: left side shows non-medicated pre-T2D vs non-T2D important species; right side shows non-medicated T2D vs non-T2D important species

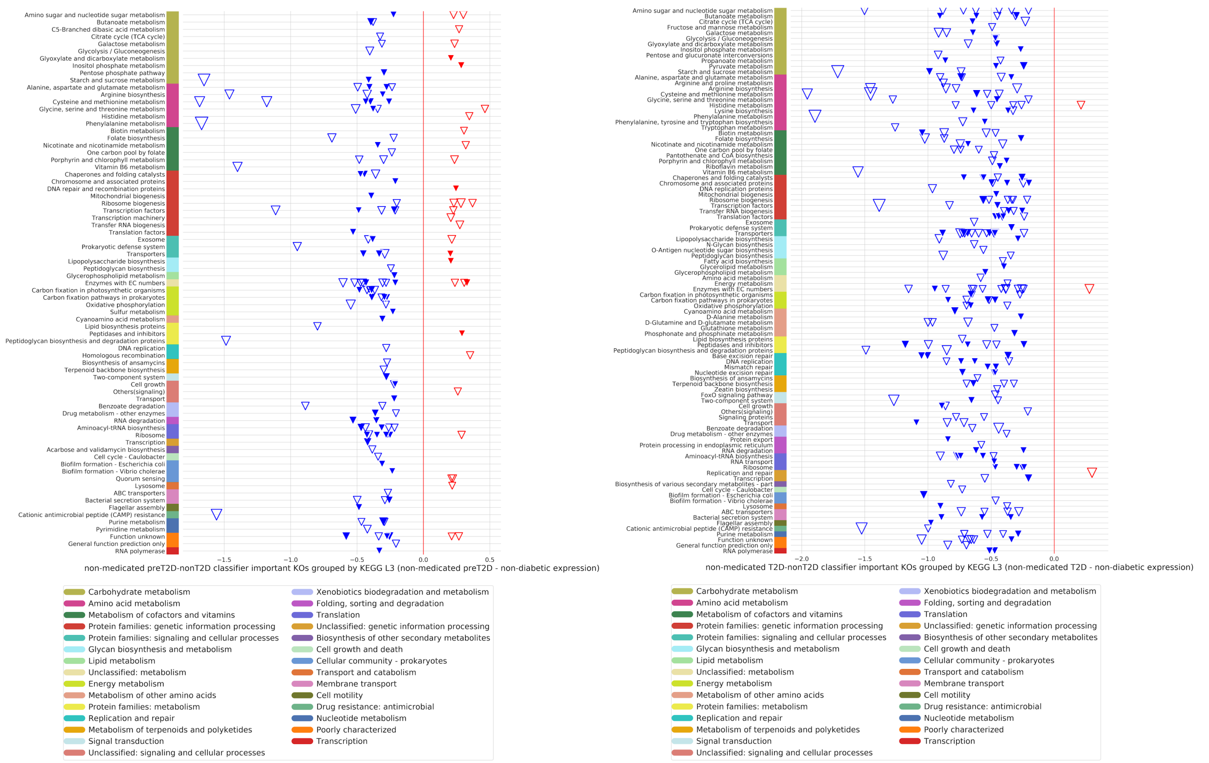


(b) Important KOs in classifiers grouped by KEGG L3 category. Left side shows non-medicated pre-T2D vs non-T2D important KOs, blue triangles show higher expression in the non-medicated T2D group, red triangles show higher expression in pre-diabetic group; right side shows non-medicated T2D vs non-T2D important KOs, blue triangles show higher expression in the non-diabetic group, red triangles show higher expression in T2D group.

Figure S6. Statistically significant richness, diversity (a-b) and feature scatter plots distinguishing T2D vs. general population using descriptive statistics (species FDR p value < 0.05; KOs FDR p value < 0.00001) (c-f). Venn diagrams representing overlapping features shared between the T2D progression and the diagnostic statistical analysis (g-j).

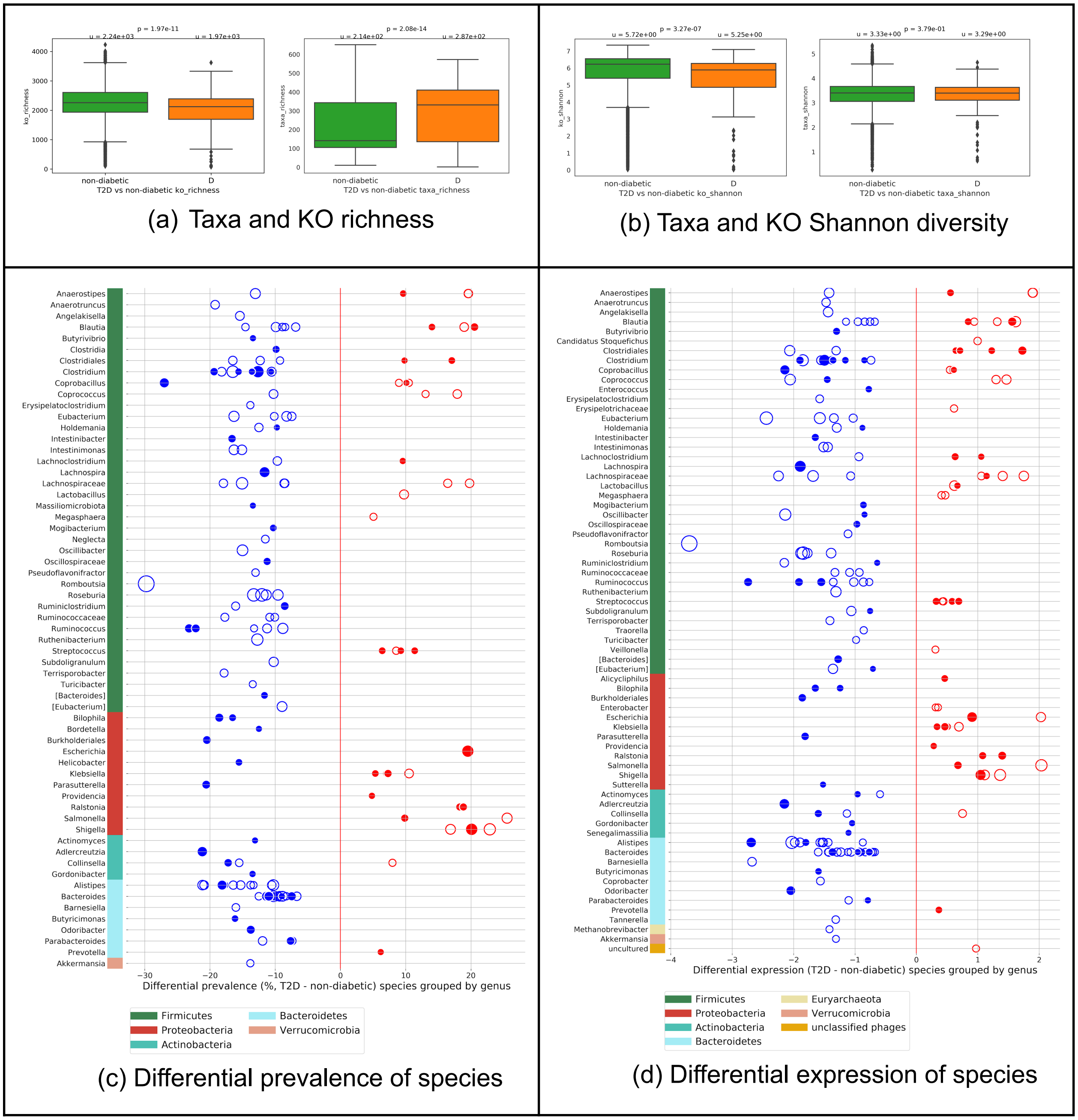

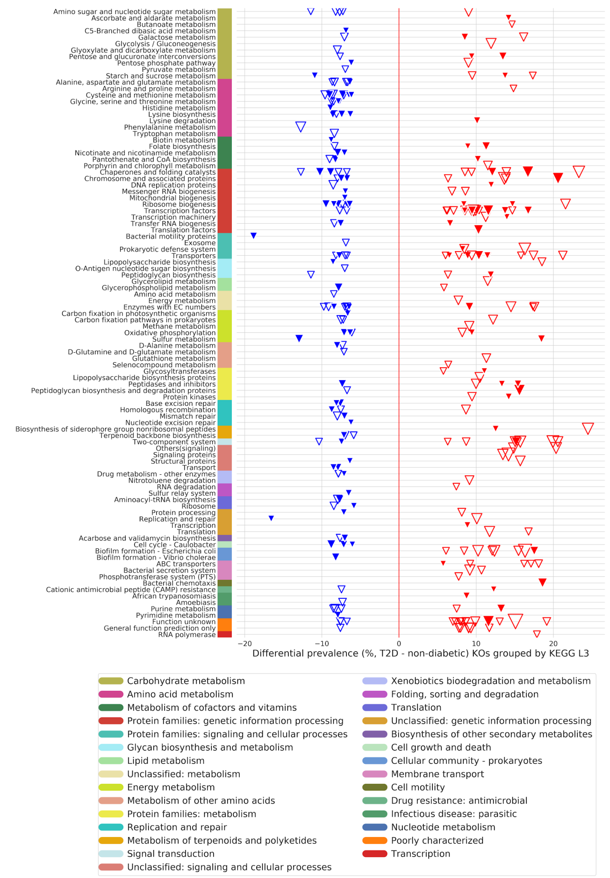


(e) Differential prevalence of KOs

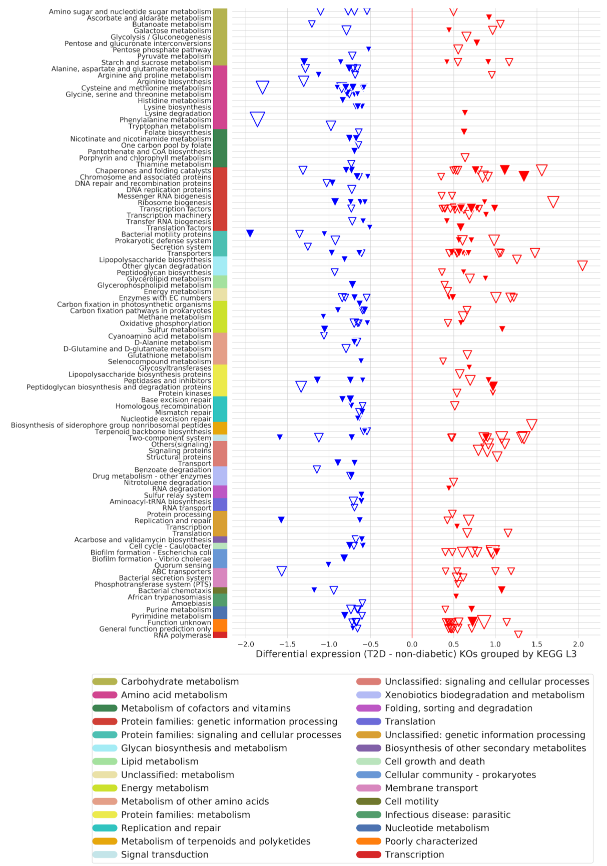


(f) Differential expression of KOs

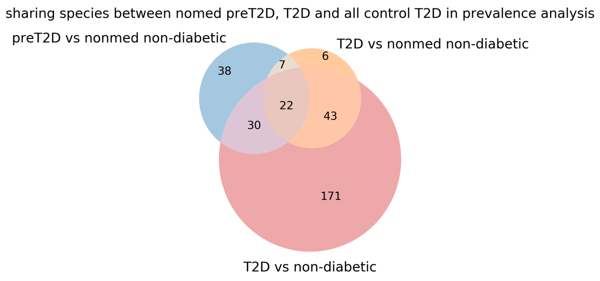


(g) Sharing significantly prevalent species between progression and diagnostic descriptive analysis

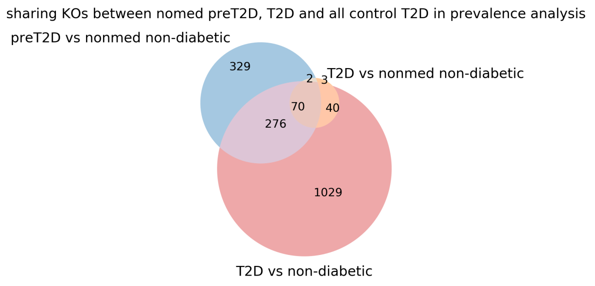


(h) Sharing significantly prevalent KOs between progression and diagnostic descriptive analysis

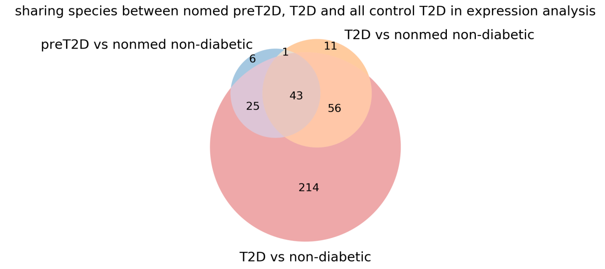


(i) Sharing significantly differentially expressed species between progression and diagnostic descriptive analysis

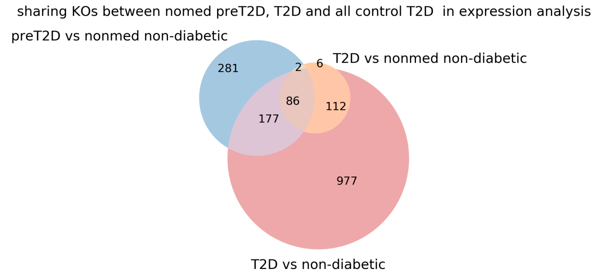


(j) Sharing significantly differentially expressed KOs between progression and diagnostic descriptive analysis

Figure S7. Important diagnostic model features using support vector machine approach (species with differential expression > 0.1; top 300 KOs with the highest coefficients and differential expression > 0.2) (a-b). c) correlation between T2D risk score and BMI in the validation cohort (n=2406)

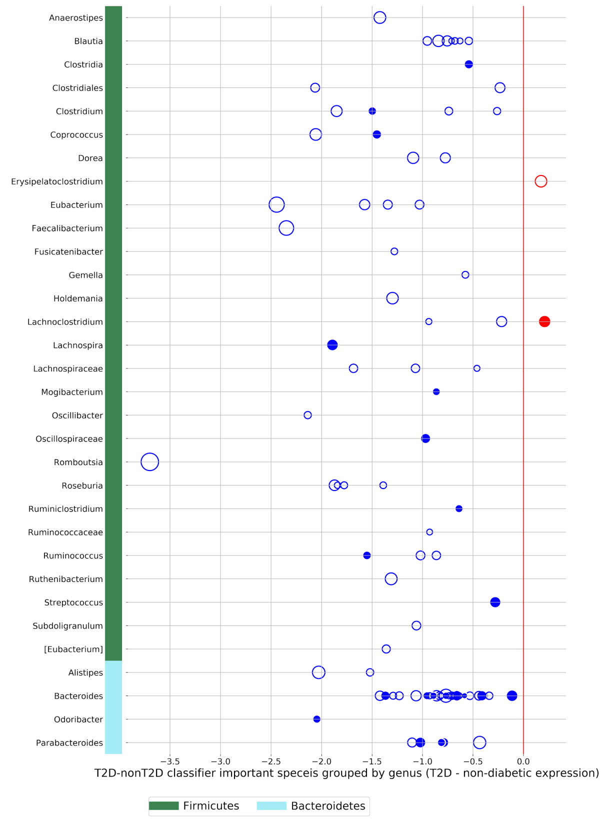


(a) Important species in T2D vs. non-diabetic classifier

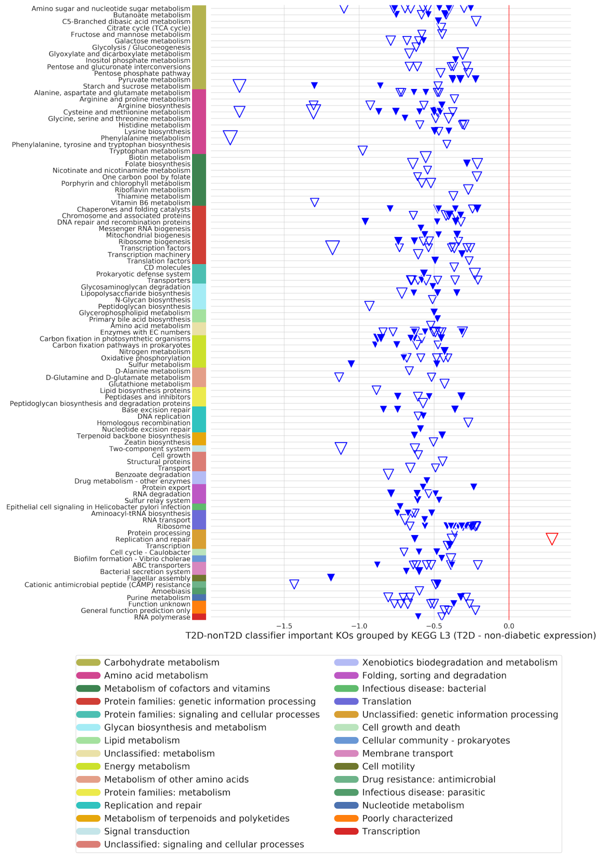


(b) Important KOs in T2D vs. non-diabetic classifier

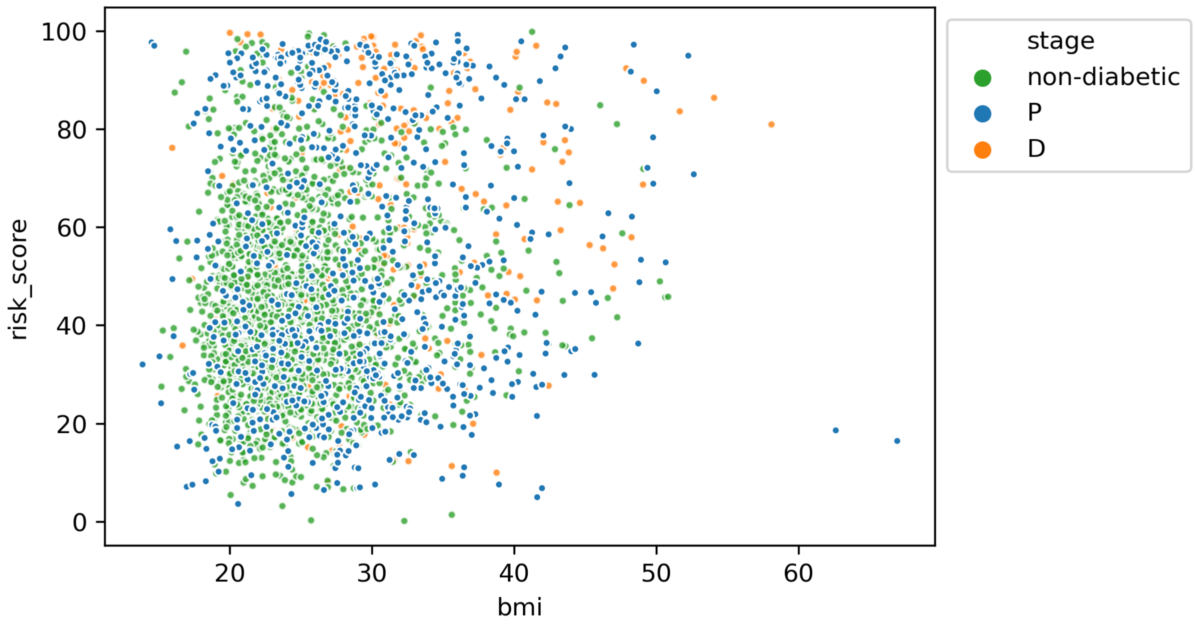


(c) Correlation between BMI and T2D risk score (rho=0.16, p=5.29e-19).

Figure S8. Richness and diversity in treatment models. (a-b) T2D metformin treated patients with good control of HbA1c (<6.5) (n=22) vs  T2D metformin treated patients who can not control their HbA1c (>=6.5)  (n=41); (c-d) T2D non-medicated responders with HbA1c <6.5 (n=29) vs  T2D non-medicated non-responders HbA1c >=6.5 (n=30).

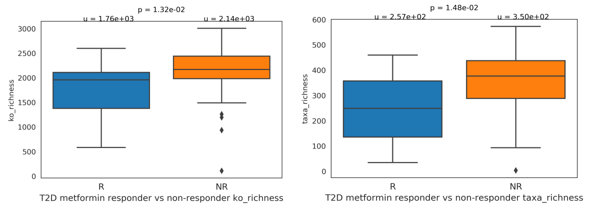


(a) Taxa and KO richness in T2D metformin responder v.s. non-responder

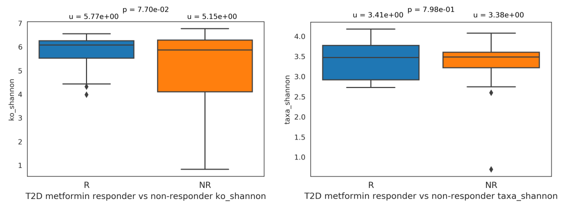


(b) Taxa and KO Shannon-diversity in T2D metformin responder v.s. non-responder

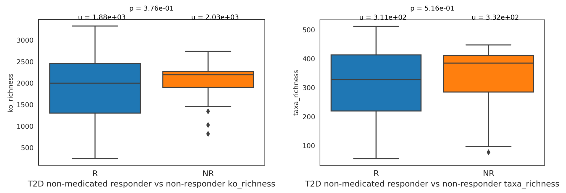


(c) Taxa and KO richness in T2D non-medicated responder v.s. non-responder

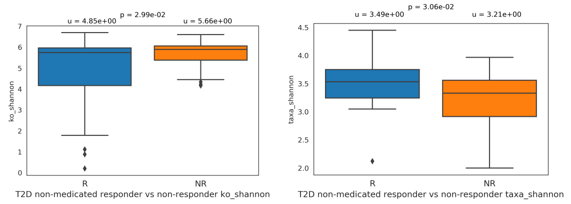


(d) Taxa and KO Shannon-diversity in T2D non-medicated responder v.s. non-responder

Figure S9. Important features in non-medicated response model using expression values and SVM approach for patients who are not using medication, but instead apply diet and lifestyle changes. Here we visualize species with differential expression > 0.1; top 300 KOs with the highest importance coefficients and differential expression > 0.2).

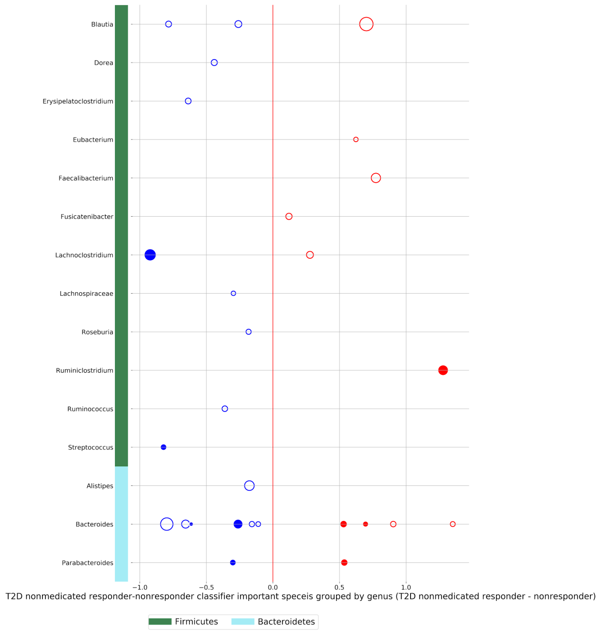


(a) Important differentially expressed species in T2D non-medicated patients who are able to control HbA1c (<=6.5) in red vs. patients who are non-medicated and not able to control HbA1c (>6.5) - in blue

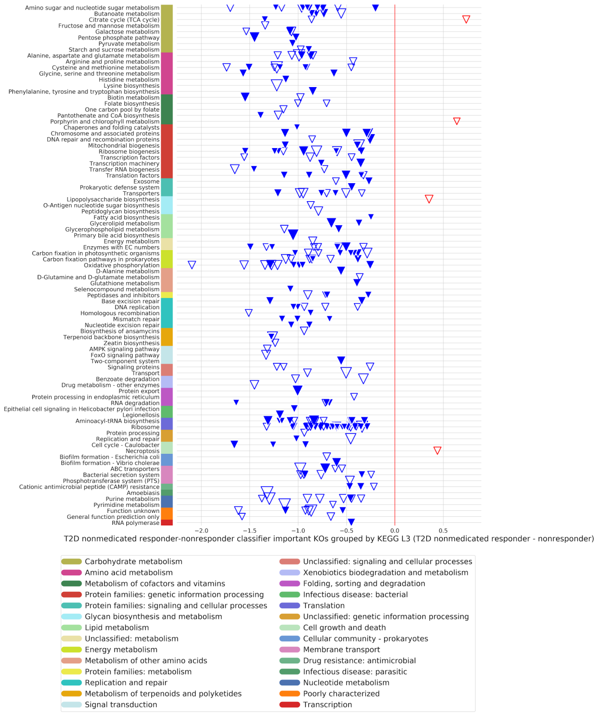


(b) Important differentially expressed KOs in T2D non-medicated patients who are able to control HbA1c (<=6.5) in red vs. patients who are non-medicated and not able to control HbA1c (>6.5) - in blue

**Taxonomical Shifts in the T2D Metatranscriptome**

The association between type 2 diabetes and an imbalanced gut microbial composition has been reported in the past many years, and studies have suggested that gut dysbiosis is a factor in the development of insulin resistance (Allin et al., 2015; Sircana et al., 2018). Dysbiosis in T2D has been characterized by a reduction of *Firmicutes* species and an increase in the typically low-abundance organisms such as *Proteobacteria* (Larsen et al., 2010). Our statistical model reveals several enriched *Firmicutes* features on both the prediabetes and T2D sides when compared to the presumed healthy population (Figure 2a, S1a, and S1b). *Firmicutes* enriched in our T2D samples include *Erysipelatoclostridium ramosum*, *Blautia producta*, and uncharacterized species from *Anaerococcus*, *Blautia*, *Clostridia*, *Clostridiales*, *Coprobacillus*, and *Streptococcus*. Although more research is required to elucidate their roles in T2D development, *Blautia* and *Streptococcus* have been positively linked to T2D-associated gut microbiota (Candela et al., 2016; Egshatyan et al., 2016). On the other hand, although the number of differentially prevalent and expressed *Proteobacteria* features is comparable in the diseased and non-diabetic cohorts, the enrichment of specific genera in the diseased populations is evident, and such distinction becomes more pronounced when progressing from the preT2D to T2D microbiome. *E. ramosum*, an opportunistic and obesogenic pathogen that may promote intestinal glucose and fat absorption (Mandić et al., 2019; Woting et al., 2014), has also been shown to be T2D-enriched in a metagenomic study of a Chinese cohort (Qin et al., 2012). Higher prevalence and abundance of *Eggerthella* and *Ralstonia* features, including *E. lenta* and *R. pickettii*, are also indicated by our statistical model in the pre-T2D and T2D samples (Figure 2a, S1a, and S1b). Interestingly, *E. lenta* is another opportunistic pathogen reported in the Chinese metagenome-wide study to be T2D-enriched, while *R. pickettii,* a pathogen capable of causing bacteremia and hospital infection outbreaks, has been suggested as an aggravator of glucose intolerance in obesity (Udayappan et al., 2017). Taken together, these results underscore the prominence of opportunistic pathogens in a disrupted microbiome and their potentials as T2D progression markers.

**Enrichment of SCFA and Succinate-Producing Taxa in Presumed Healthy Cohort**

Studies have demonstrated the multi-mechanistic role of short-chain fatty acids in contributing to intestinal homeostasis. In particular, butyrate and acetate have been studied for their beneficial effects on insulin secretion, energy metabolism, and glucose tolerance (Gao et al., 2009; De Vadder et al., 2014; Yamashita et al., 2007). Our descriptive analysis identifies several butyrate producers such as *Butyricimonas*, *Butyrivibrio*, *Coprococcus*, and *Odoribacter* to be differentially expressed in the non-diabetic samples (Figure 2a & S1b). The enrichment of acetate producers such as *Alistipes*, *Ruminococcus*, *Odoribacter*, and *Paraprevotella* in the non-diabetic cohort is also noted. The presence of these SCFA-producing species aligns well with previous findings, highlighting their significance in normal or improved metabolic health. In addition to the enrichment of SCFA-producing species, the descriptive model suggests the capacity to produce succinate, a precursor/intermediate for SCFA, as a distinguishing feature, as many succinate-producing bacteria are present at higher abundance in the non-diabetic population compared to either T2D or preT2D (Figure 2a, S1a and S1b). The observed taxa classify under the phylum *Bacteroidetes*, which is known to harbor numerous succinate-producing species, and include *Alistipes putredinis*, *Alistipes ihumii*, *Parabacteroides*, *Paraprevotella clara*, *Odoribacter splanchnicus*, and *Ruminococcus callidus* (Rautio et al., 2003; Parker et al., 2020; Sakamoto & Benno, 2006; Morotomi et al., 2009; Hiippala et al., 2020; La Reau & Suen, 2018). While the linkage between succinate, dysbiosis, and inflammatory diseases such as IBD is recognized, succinate has also been suggested to have beneficial metabolic effects and play a role in glucose tolerance and insulin sensitivity (De Vadder et al., 2016; Fernández-Veledo & Vendrell, 2019). Microbiota-derived succinate has been shown to promote glucose homeostasis by serving as a substrate of intestinal gluconeogenesis (IGN), leading to the inhibition of hepatic glucose production (De Vadder et al., 2016). Additionally, esters of succinic acid have been shown to be metabolized into succinate intracellularly and exert insulinotropic effects (MacDonald & Fahien, 1988).

**Bile-Resistant Bacteria Associated with Presumed Healthy Individuals**

Colonic bacteria are capable of modifying and deconjugating primary bile acids synthesized in hepatocytes into secondary bile acids, resulting in a pool of diversified bile forms in the intestines. The relationship between the chemical diversity of the bile acids, their signaling properties, and bile homeostasis in T2D patients has been demonstrated increasingly. From our descriptive analysis, the differential expression of bile-resistant microbes, such as *Alistipes*, *Bilophila*, *Bacteroides*, and *Sutterella* is observed in the presumed healthy cohort (Figure 2a & S1a, and S1b). The abundance of *Alistipes*, *Bilophila* and *Bacteroides* has been shown to increase in animal-based diets, which is presumably the result of a higher fat intake and therefore increased activity of bile secretion compared to plant-based diets (David et al., 2014). Two *Bilophila* features are consistently enriched on the expression and prevalence level in the non-diabetes population with respect to T2D patients (Figure 2a and S1a). Notably, *Bilophila wadsworthia* utilizes taurine as an electron acceptor and its growth has been shown to be promoted by high-fat diets due to the increased availability of taurine-conjugated bile acids synthesized to facilitate lipid digestion (Devkota et al., 2012). Functionally, although the statistical model does not pinpoint any KOs in the pathways of secondary bile acid production, our machine learning model identifies bile salt hydrolase (K01442) to be an important feature for the prediction of preT2D and T2D microbiome. Overall, the enrichment of bile-resistant bacteria may be the result of unrestricted diet in the presumed healthy cohort, altered bile acid homeostasis in T2D patients, or other feedback mechanisms underlying the T2D microbiome.

**Taxa Features in the Metformin Treatment Model**

Mounting evidence over the past several years suggests metformin as a major modifier of the gut microbiota, and metformin treatments have been shown to promote the growth of certain intestinal bacteria, notably *Akkermansia muciniphila* and *Escherichia spp*. (Elbere et al., 2018; Karlsson et al., 2013; Lee & Ko, 2014; Wu et al., 2017). Our treatment classifiers present a different view by predicting microbes potentially important in discriminating between those who can and cannot control their HbA1c in response to metformin treatment (Figure 4). In general, the model predicts the importance of several *Bacteroides* and *Parabacteroides* species, and these genera have been shown to be among the most frequently altered groups in metformin treatments (Huang et al., 2020). *Bacteroides thetaiotaomicron*, an opportunistic pathogen and common inhabitant of a normal gut microbiome, is capable of fermenting a broad range of polysaccharides and considered by the model to be important in predicting those who cannot control their HbA1c levels. *B. thetaiotaomicron* has been shown to modulate inflammation by activating the Treg pathways, which play a role in reducing inflammatory response and improving insulin resistance (Hoffmann et al., 2016), but its ability to enhance the energy harvest and nutrient absorption of the host has also been demonstrated (Hooper et al., 2001; Samuel & Gordon, 2006). Similarly, *B. eggerthii* is identified as a potential predictor of non-responders. This microbe has been shown to increase in abundance in T2D microbiome and to aggravate colitis in mice models (Dziarski et al., 2016; Medina-Vera et al., 2019). Overall, we find that more research is needed to understand the taxa in both the responder and non-responder groups, as the features seem to present a complex picture of metformin efficacy.

### REFERENCES


Allin, K. H., Nielsen, T., & Pedersen, O. (2015). Mechanisms in endocrinology: Gut microbiota in patients with type 2 diabetes mellitus. *European Journal of Endocrinology*, *172*(4), R167-177. https://doi.org/10.1530/EJE-14-0874

Candela, M., Biagi, E., Soverini, M., Consolandi, C., Quercia, S., Severgnini, M., Peano, C., Turroni, S., Rampelli, S., Pozzilli, P., Pianesi, M., Fallucca, F., & Brigidi, P. (2016). Modulation of gut microbiota dysbioses in type 2 diabetic patients by macrobiotic Ma-Pi 2 diet. *The British Journal of Nutrition*, *116*(1), 80–93. https://doi.org/10.1017/S0007114516001045

David, L. A., Maurice, C. F., Carmody, R. N., Gootenberg, D. B., Button, J. E., Wolfe, B. E., Ling, A. V., Devlin, A. S., Varma, Y., Fischbach, M. A., Biddinger, S. B., Dutton, R. J., & Turnbaugh, P. J. (2014). Diet rapidly and reproducibly alters the human gut microbiome. *Nature*, *505*(7484), 559–563. https://doi.org/10.1038/nature12820

De Vadder, F., Kovatcheva-Datchary, P., Zitoun, C., Duchampt, A., Bäckhed, F., & Mithieux, G. (2016). Microbiota-Produced Succinate Improves Glucose Homeostasis via Intestinal Gluconeogenesis. *Cell Metabolism*, *24*(1), 151–157. https://doi.org/10.1016/j.cmet.2016.06.013

De Vadder, F., Kovatcheva-Datchary, P., Goncalves, D., Vinera, J., Zitoun, C., Duchampt, A., Bäckhed, F., & Mithieux, G. (2014). Microbiota-Generated Metabolites Promote Metabolic Benefits via Gut-Brain Neural Circuits. *Cell*, *156*(1), 84–96. https://doi.org/10.1016/j.cell.2013.12.016

Devkota, S., Wang, Y., Musch, M. W., Leone, V., Fehlner-Peach, H., Nadimpalli, A., Antonopoulos, D. A., Jabri, B., & Chang, E. B. (2012). Dietary-fat-induced taurocholic acid promotes pathobiont expansion and colitis in Il10-/- mice. *Nature*, *487*(7405), 104–108. https://doi.org/10.1038/nature11225

Dziarski, R., Park, S. Y., Kashyap, D. R., Dowd, S. E., & Gupta, D. (2016). Pglyrp-Regulated Gut Microflora Prevotella falsenii, Parabacteroides distasonis and Bacteroides eggerthii Enhance and Alistipes finegoldii Attenuates Colitis in Mice. *PloS One*, *11*(1), e0146162. https://doi.org/10.1371/journal.pone.0146162

Egshatyan, L., Kashtanova, D., Popenko, A., Tkacheva, O., Tyakht, A., Alexeev, D., Karamnova, N., Kostryukova, E., Babenko, V., Vakhitova, M., & Boytsov, S. (2016). Gut microbiota and diet in patients with different glucose tolerance. *Endocrine Connections*, *5*(1), 1–9. https://doi.org/10.1530/EC-15-0094

Elbere, I., Kalnina, I., Silamikelis, I., Konrade, I., Zaharenko, L., Sekace, K., Radovica-Spalvina, I., Fridmanis, D., Gudra, D., Pirags, V., & Klovins, J. (2018). Association of metformin administration with gut microbiome dysbiosis in healthy volunteers. *PLoS ONE*, *13*(9). https://doi.org/10.1371/journal.pone.0204317

Fernández-Veledo, S., & Vendrell, J. (2019). Gut microbiota-derived succinate: Friend or foe in human metabolic diseases? *Reviews in Endocrine & Metabolic Disorders*, *20*(4), 439–447. https://doi.org/10.1007/s11154-019-09513-z

Gao, Z., Yin, J., Zhang, J., Ward, R. E., Martin, R. J., Lefevre, M., Cefalu, W. T., & Ye, J. (2009). Butyrate Improves Insulin Sensitivity and Increases Energy Expenditure in Mice. *Diabetes*, *58*(7), 1509–1517. https://doi.org/10.2337/db08-1637

Hiippala, K., Barreto, G., Burrello, C., Diaz-Basabe, A., Suutarinen, M., Kainulainen, V., Bowers, J. R., Lemmer, D., Engelthaler, D. M., Eklund, K. K., Facciotti, F., & Satokari, R. (2020). Novel Odoribacter splanchnicus Strain and Its Outer Membrane Vesicles Exert Immunoregulatory Effects in vitro. *Frontiers in Microbiology*, *11*, 575455. https://doi.org/10.3389/fmicb.2020.575455

Hoffmann, T. W., Pham, H.-P., Bridonneau, C., Aubry, C., Lamas, B., Martin-Gallausiaux, C., Moroldo, M., Rainteau, D., Lapaque, N., Six, A., Richard, M. L., Fargier, E., Le Guern, M.-E., Langella, P., & Sokol, H. (2016). Microorganisms linked to inflammatory bowel disease-associated dysbiosis differentially impact host physiology in gnotobiotic mice. *The ISME Journal*, *10*(2), 460–477. https://doi.org/10.1038/ismej.2015.127

Hooper, L. V., Wong, M. H., Thelin, A., Hansson, L., Falk, P. G., & Gordon, J. I. (2001). Molecular analysis of commensal host-microbial relationships in the intestine. *Science (New York, N.Y.)*, *291*(5505), 881–884. https://doi.org/10.1126/science.291.5505.881

Huang, X., Hong, X., Wang, J., Sun, T., Yu, T., Yu, Y., Fang, J., & Xiong, H. (2020). Metformin elicits antitumour effect by modulation of the gut microbiota and rescues Fusobacterium nucleatum-induced colorectal tumourigenesis. *EBioMedicine*, *61*, 103037. https://doi.org/10.1016/j.ebiom.2020.103037

Karlsson, F. H., Tremaroli, V., Nookaew, I., Bergström, G., Behre, C. J., Fagerberg, B., Nielsen, J., & Bäckhed, F. (2013). Gut metagenome in European women with normal, impaired and diabetic glucose control. *Nature*, *498*(7452), 99–103. https://doi.org/10.1038/nature12198

La Reau, A. J., & Suen, G. (2018). The Ruminococci: Key symbionts of the gut ecosystem. *Journal of Microbiology (Seoul, Korea)*, *56*(3), 199–208. https://doi.org/10.1007/s12275-018-8024-4

Larsen, N., Vogensen, F. K., van den Berg, F. W. J., Nielsen, D. S., Andreasen, A. S., Pedersen, B. K., Al-Soud, W. A., Sørensen, S. J., Hansen, L. H., & Jakobsen, M. (2010). Gut microbiota in human adults with type 2 diabetes differs from non-diabetic adults. *PloS One*, *5*(2), e9085. https://doi.org/10.1371/journal.pone.0009085

Lee, H., & Ko, G. (2014). Effect of metformin on metabolic improvement and gut microbiota. *Applied and*

*Environmental Microbiology*, *80*(19), 5935–5943. https://doi.org/10.1128/AEM.01357-14
MacDonald, M. J., & Fahien, L. A. (1988). Glyceraldehyde phosphate and methyl esters of succinic acid. Two “new” potent insulin secretagogues. *Diabetes*, *37*(7), 997–999. https://doi.org/10.2337/diab.37.7.997
Mandić, A. D., Woting, A., Jaenicke, T., Sander, A., Sabrowski, W., Rolle-Kampcyk, U., von Bergen, M., & Blaut, M. (2019). Clostridium ramosum regulates enterochromaffin cell development and serotonin release. *Scientific Reports*, *9*(1), 1177. https://doi.org/10.1038/s41598-018-38018-z
Medina-Vera, I., Sanchez-Tapia, M., Noriega-López, L., Granados-Portillo, O., Guevara-Cruz, M., Flores-López, A., Avila-Nava, A., Fernández, M. L., Tovar, A. R., & Torres, N. (2019). A dietary intervention with functional foods reduces metabolic endotoxaemia and attenuates biochemical abnormalities by modifying faecal microbiota in people with type 2 diabetes. *Diabetes & Metabolism*, *45*(2), 122–131. https://doi.org/10.1016/j.diabet.2018.09.004
Morotomi, M., Nagai, F., Sakon, H., & Tanaka, R. (2009). Paraprevotella clara gen. Nov., sp. Nov. And Paraprevotella xylaniphila sp. Nov., members of the family “Prevotellaceae” isolated from human faeces. *International Journal of Systematic and Evolutionary Microbiology*, *59*(Pt 8), 1895–1900. https://doi.org/10.1099/ijs.0.008169-0
Parker, B. J., Wearsch, P. A., Veloo, A. C. M., & Rodriguez-Palacios, A. (2020). The Genus Alistipes: Gut Bacteria With Emerging Implications to Inflammation, Cancer, and Mental Health. *Frontiers in Immunology*, *11*, 906. https://doi.org/10.3389/fimmu.2020.00906
Qin, J., Li, Y., Cai, Z., Li, S., Zhu, J., Zhang, F., Liang, S., Zhang, W., Guan, Y., Shen, D., Peng, Y., Zhang, D., Jie, Z., Wu, W., Qin, Y., Xue, W., Li, J., Han, L., Lu, D., … Wang, J. (2012). A metagenome-wide association study of gut microbiota in type 2 diabetes. *Nature*, *490*(7418), 55–60. https://doi.org/10.1038/nature11450
Rautio, M., Eerola, E., Väisänen-Tunkelrott, M.-L., Molitoris, D., Lawson, P., Collins, M. D., & Jousimies-Somer, H. (2003). Reclassification of Bacteroides putredinis (Weinberg et al., 1937) in a new genus Alistipes gen. Nov., as Alistipes putredinis comb. Nov., and description of Alistipes finegoldii sp. Nov., from human sources. *Systematic and Applied Microbiology*, *26*(2), 182–188. https://doi.org/10.1078/072320203322346029
Sakamoto, M., & Benno, Y. (2006). Reclassification of Bacteroides distasonis, Bacteroides goldsteinii and Bacteroides merdae as Parabacteroides distasonis gen. Nov., comb. Nov., Parabacteroides goldsteinii comb. Nov. And Parabacteroides merdae comb. Nov. *International Journal of Systematic and Evolutionary Microbiology*, *56*(Pt 7), 1599–1605. https://doi.org/10.1099/ijs.0.64192-0
Samuel, B. S., & Gordon, J. I. (2006). A humanized gnotobiotic mouse model of host-archaeal-bacterial mutualism. *Proceedings of the National Academy of Sciences of the United States of America*, *103*(26), 10011–10016. https://doi.org/10.1073/pnas.0602187103
Sircana, A., Framarin, L., Leone, N., Berrutti, M., Castellino, F., Parente, R., De Michieli, F., Paschetta, E., & Musso, G. (2018). Altered Gut Microbiota in Type 2 Diabetes: Just a Coincidence? *Current Diabetes Reports*, *18*(10), 98. https://doi.org/10.1007/s11892-018-1057-6
Udayappan, S. D., Kovatcheva-Datchary, P., Bakker, G. J., Havik, S. R., Herrema, H., Cani, P. D., Bouter, K. E., Belzer, C., Witjes, J. J., Vrieze, A., de Sonnaville, E. S. V., Chaplin, A., van Raalte, D. H., Aalvink, S., Dallinga-Thie, G. M., Heilig, H. G. H. J., Bergström, G., van der Meij, S., van Wagensveld, B. A., … Nieuwdorp, M. (2017). Intestinal Ralstonia pickettii augments glucose intolerance in obesity. *PloS One*, *12*(11), e0181693. https://doi.org/10.1371/journal.pone.0181693
Woting, A., Pfeiffer, N., Loh, G., Klaus, S., & Blaut, M. (2014). Clostridium ramosum promotes high-fat diet-induced obesity in gnotobiotic mouse models. *MBio*, *5*(5), e01530-01514. https://doi.org/10.1128/mBio.01530-14
Wu, H., Esteve, E., Tremaroli, V., Khan, M. T., Caesar, R., Mannerås-Holm, L., Ståhlman, M., Olsson, L. M., Serino, M., Planas-Fèlix, M., Xifra, G., Mercader, J. M., Torrents, D., Burcelin, R., Ricart, W., Perkins, R., Fernàndez-Real, J. M., & Bäckhed, F. (2017). Metformin alters the gut microbiome of individuals with treatment-naive type 2 diabetes, contributing to the therapeutic effects of the drug. *Nature Medicine*, *23*(7), 850–858. https://doi.org/10.1038/nm.4345
Yamashita, H., FUJISAWA, K., ITO, E., IDEI, S., KAWAGUCHI, N., KIMOTO, M., HIEMORI, M., & TSUJI, H. (2007). Improvement of Obesity and Glucose Tolerance by Acetate in Type 2 Diabetic Otsuka Long-Evans Tokushima Fatty (OLETF) Rats. *Bioscience, Biotechnology, and Biochemistry*, *71*(5), 1236–1243. https://doi.org/10.1271/bbb.60668
